# A Blockchain-Enabled Federated Ensemble Framework for Privacy-Preserving Mortality Prediction Across Heterogeneous Healthcare Systems

**DOI:** 10.64898/2026.11.19.26368466

**Authors:** Mostak Ahmed, Sabera Tabassum Wahid, Golam Sarwar Khondoker, Golam Rabbi, Rony Md Ariful Islam, Md Mostafizur Rahman, Al Rafy, Sanjida Islam, Md. Masuqul Haque

**Author notes:** Corresponding author: Mostak Ahmed, Information and Communication Technology (ICT) Cell, University of Dhaka, Dhaka 1000, Bangladesh.

## Abstract

**Objective:** To test whether a smart contract that admits hospital-trained models to a shared mortality model only after peer hospitals verify their performance can exclude poisoned or substituted models, and at what cost.

**Design:** Retrospective simulation study of a federated learning protocol.

**Methodology:** In the Blockchain-Enabled Federated Ensemble (BFE), hospitals committed SHA-256 digests of locally trained XGBoost and random-forest models to a Solidity contract; peers posted digest-bound validation AUROCs, and the contract admitted models with a median peer AUROC of at least 0.70. Admitted models were pooled by weighted logit averaging (BFE-PW) or stacking (BFE-Stack) with federated recalibration. Code and drafts were produced with an AI coding assistant under the authors’ supervision.

**Setting:** Four hospitals simulated from one academic centre with age, renal case-mix and laboratory-availability skew; a data-scarce regime keeping 5–20% of each hospital’s training and validation patients; and a natural partition by care era.

**Participants:** 425,215 adult admissions (186,103 patients; 2.35% in-hospital mortality) in MIMIC-IV v3.1, with predictors from the first 24 hours.

**Interventions:** BFE against centralised, local-only, FedAvg, FedProx, median and Multi-Krum FedAvg and federated logistic regression, under seven attacks on training, attestation, recalibration and model delivery.

**Main outcome measures:** AUROC and AUPRC (joint patient-clustered bootstrap), calibration, attack effects, gas and transactions.

**Results:** Under gross label flipping, FedAvg lost 29% of its AUPRC while the contract excluded the poisoned models in every seed (Multi-Krum FedAvg was equally protected); all substituted models were detected, and collusion blocked release. A targeted flip of female deaths passed the gate and lowered sensitivity for deaths in women from 32.8% to 27.5%; poisoned recalibration inverted BFE-PW (AUROC 0.098) unless a median-of-sites recalibration was used (0.902). With large hospitals, BFE-PW matched FedAvg in AUROC (0.902) and BFE-Stack matched FedProx (0.907 and 0.906), below local-only (0.910) and centralised XGBoost (0.915), with lower AUPRC; with 5% of the data, BFE-PW exceeded local models (0.866 against 0.844). Pooled calibration hid two-fold under-prediction at one hospital (observed-to-expected ratio 2.13), which a local intercept corrected (0.93). One aggregation used 37 transactions and 3.3 million gas, rising roughly quadratically with consortium size.

**Conclusions:** Contract-enforced peer attestation gave auditable, automatic exclusion of grossly poisoned and substituted models at a small accuracy cost, but did not detect attacks that preserve validation discrimination. It is most useful for consortia of small hospitals that need a verifiable record of which models entered a shared tool.

**Plain language summary:** Hospitals can build better risk-prediction tools by learning from each other’s patients, but privacy rules make pooling records hard. Federated learning lets each hospital train on its own data and share only the model. A hospital could, however, contribute a bad model by error or on purpose, and no one can later check which models went into the shared tool. In our design each hospital records a digital fingerprint of its model on a blockchain, other hospitals test the model and record their scores, and a program on the blockchain lets the model in only if it scored well enough. We simulated groups of hospitals using 425,215 hospital stays.

**Key findings:**

- The shared tool was almost as accurate as standard federated learning, and better than each hospital alone when hospitals were small.
- Models trained on completely falsified outcomes were always kept out, and swapped models were always caught.
- Subtler sabotage got through: hiding deaths in women made the tool miss more deaths in women, and falsifying the final adjustment step could reverse its predictions unless a robust adjustment was used.
- At one hospital that measured fewer laboratory tests, the shared tool underestimated risk by about half; a simple local adjustment, made by that hospital alone, fixed it.
- Recording everything on a blockchain had a measurable cost that grows quickly as more hospitals join.

The design makes the shared tool auditable, but it must be tested next on real data from separate hospitals.

## 1. Introduction

Federated learning (FL) lets hospitals build a shared prediction model without pooling patient records, which data-protection law such as HIPAA and the GDPR restricts [1–4], and it can approach centralised accuracy for clinical out-comes [5–8]. Two obstacles remain for the tabular electronic health record (EHR) data behind most hospital risk models. First, gradient-boosted and random-forest ensembles often outperform deep networks on tabular data [9,10], yet they have no shared parameter space and cannot be combined by FedAvg-style averaging. Second, a consortium of independent hospitals needs a verifiable record of which contributed models entered the shared model, on what evidence, and whether the deployed artefact is the one that was validated. Without it, one poisoned or swapped contribution can degrade the shared model unnoticed [11,12], and robust aggregation rules designed for neural-network updates protect only partly [13,14].

Hospitals also differ in patient age, case mix and the laboratory tests they order, so their data are not independent and identically distributed (non-IID). Under such skew, iterative parameter averaging can drift and converge slowly [15–18], and the clinical remedies proposed so far [17,19–21] assume a gradient-trained model. Tree ensembles can be federated without averaging, by lossless federated boosting, which exchanges encrypted gradient statistics at every split [22], or by one-shot ensembling of locally trained models [23–25].

We therefore ask whether peer-attested admission of whole, locally trained tree ensembles, with every attestation and decision recorded on a smart contract, can give a consortium that verifiable record at a small cost in accuracy, and when collaboration through such a protocol pays off.

Ledger-based clinical FL already records updates, consent and site-level misconduct for neural or linear models [26–31] (Section 2). We found no clinical study in which a contract admits ensemble members by a published rule over hash-bound peer validation scores, and none that tested post-validation artefact substitution and collusion separately from training-time poisoning. The Blockchain-Enabled Federated Ensemble (BFE) studied here federates tree ensembles in one exchange and decides admission per committed artefact, and any party can recompute each decision from on-chain attestations.

We present BFE as a governance protocol for federating tree ensembles, and evaluate it in a simulation study on 425,215 adult admissions from MIMIC-IV v3.1 [32,33]. Our contributions are:

1. **A governance protocol for one-shot federation of tree ensembles**. Hospitals commit SHA-256 digests of their XGBoost and random-forest members to a smart contract. Peers post validation AUROCs (area under the receiver operating characteristic curve) that must name the committed digest, and the contract admits a member whose median peer AUROC is at least 0.70. The admitted set and weights can be recomputed from the ledger, so a deviating aggregator is detectable, though not prevented; recalibration and stacking coefficients are committed only by digest. The protocol extends one-shot ensembling [23,24] by giving member selection to a contract that applies a published rule to peer evidence.
2. **Accuracy and calibration in simulated consortia**. Four hospitals were simulated by rule-based age, renal and laboratory-availability skew, with an IID control, and MIMIC-IV care eras (anchor-year groups 2008–10, 2011–13, 2014–16, 2017–22) gave a second, non-simulated partition. With large hospitals BFE matched FedAvg or FedProx in AUROC but stayed below local-only and centralised XGBoost, and when each hospital kept only 5% of its patients it exceeded local models, as FedAvg did. Comparators include federated logistic regression; the primary inference is a joint patient-clustered bootstrap.
3. **A threat evaluation that separates attack surfaces**: full, partial and targeted label flips, poisoned recalibration and threshold steps, false attestation, post-validation artefact substitution and collusion by half of the consortium, against an oracle ensemble and robust FedAvg [13,14]. It shows which surfaces the contract closes (gross poisoning by the gate, substitution by the digest check), where it fails closed (collusion: no model is released) and which it leaves open (targeted and recalibration poisoning): a gate certifies average discrimination, not subgroup safety or calibration.
4. **Site-level calibration repair and ledger cost**. A local site-specific intercept, which shares no data, corrected two-fold under-prediction at the hospital with a restricted laboratory menu (observed-to-expected ratio, O/E, 2.13 to 0.93), whereas a robust median-of-sites recalibration did not. Gas and latency are reported for 4 to 32 hospitals.

## 2. Related Work

### 2.1 Mortality prediction on tabular EHR

MIMIC-III [34], MIMIC-IV [32] and the multi-centre eICU database [35] made in-hospital and intensive care unit (ICU) mortality a standard prediction task, and EHR-based models exceed conventional severity scores [36–38]. On tabular features, tree ensembles often outperform deep networks [9,10]. Performance falls when models are transported between units or institutions [39], and reporting guidance asks for discrimination, calibration and clinical utility [40,41].

### 2.2 Federated learning under clinical heterogeneity

Clinical FL has ranged from sparse linear models [42] and patient clustering on eICU [43] to FedAvg-style mortality and oxygen-requirement prediction across hospitals [6,44]. Fe-dAvg’s convergence worsens with heterogeneity [15,16], and remedies include proximal terms [17] and re-weighting or decoupling schemes [19–21]. Across real sites, heterogene-ity arises from demographics, medication use and site practice [45], and federated XGBoost across five ICU databases trailed centralised training [46,47]. Reviews find decentralised models comparable to centralised ones in AUROC and better than local models [8,48], but hospitals with large datasets gain little [49,50].

### 2.3 Federating tree models and one-shot ensembles

Lossless federated boosting exchanges encrypted gradient statistics at every split [22], SimFL cuts that cost with locality-sensitive hashing [51], and FedXGBllr sends local ensembles once and learns their combination [52]. One-shot FL ensembles locally trained models after a single round, for example admitting a model that reaches a baseline AUROC on its own site’s data [23]; FENS adds a federatedly trained aggregator [24], the analogue of BFE-Stack. Federated random forests improved on local models [53], helped small hospitals most [49] and were evaluated across eleven Austrian hospitals [25]. On MIMIC-IV split into five non-IID sites, inference-time ensembling recovered less area under the precision–recall curve (AUPRC) than FedAvg [54]. None of these studies screens contributions against adversaries; BFE keeps the one-shot structure and adds peer-attested admission executed by a contract.

### 2.4 Poisoning defences and verifiable contribution screening

Byzantine-robust aggregation bounds the influence of outlying updates by Krum and Multi-Krum [13], the coordinate-wise median and trimmed mean [14] or a server-held root dataset [55], and ensembles of FL models give certified robustness to a few malicious clients [56]. On ledgers, committees validate updates by Multi-Krum, consensus or voting [57–59]. In health, Kalapaaking et al. verified models by secure multi-party computation before blockchain aggregation [60], FedSecure-Chain logged client trust on a ledger after anomaly detection and robust aggregation [61], and blockchain FL for retinal imaging resisted label flipping [62]. Kuo and colleagues screened submitted clinical models with auditing, coefficient and performance detectors [28] and, on a blockchain network, quarantined sites only when a misbehaviour budget ran out, to tolerate false positives [29]. These schemes screen updates or sites, using detectors, a committee or the server.

### 2.5 Blockchain for clinical model governance

Ledgers were proposed early for biomedical data provenance [63]. ModelChain proposed privacy-preserving predictive modelling on a private blockchain [64], GloreChain fitted logistic regression on one [65], and QuorumChain and WebQuorumChain used smart contracts and a proof-of-quorum protocol to keep clinical model learning going when sites drop out [30,31]. FedEnTrust used smart contracts for registration and role-based access over a distilled federated ensemble for diabetes prediction [66], and BlockFedMed governed consent, integrity and incentives for ICU mortality prediction [27]. Outside medicine, DFedForest built random forests from decision trees shared through a blockchain [67]. Reviews describe a growing field [68], yet a systematic review found only 11 implemented and quantitatively evaluated biomedical blockchain systems [69]. Table 1 compares the closest designs.

**Table 1.**
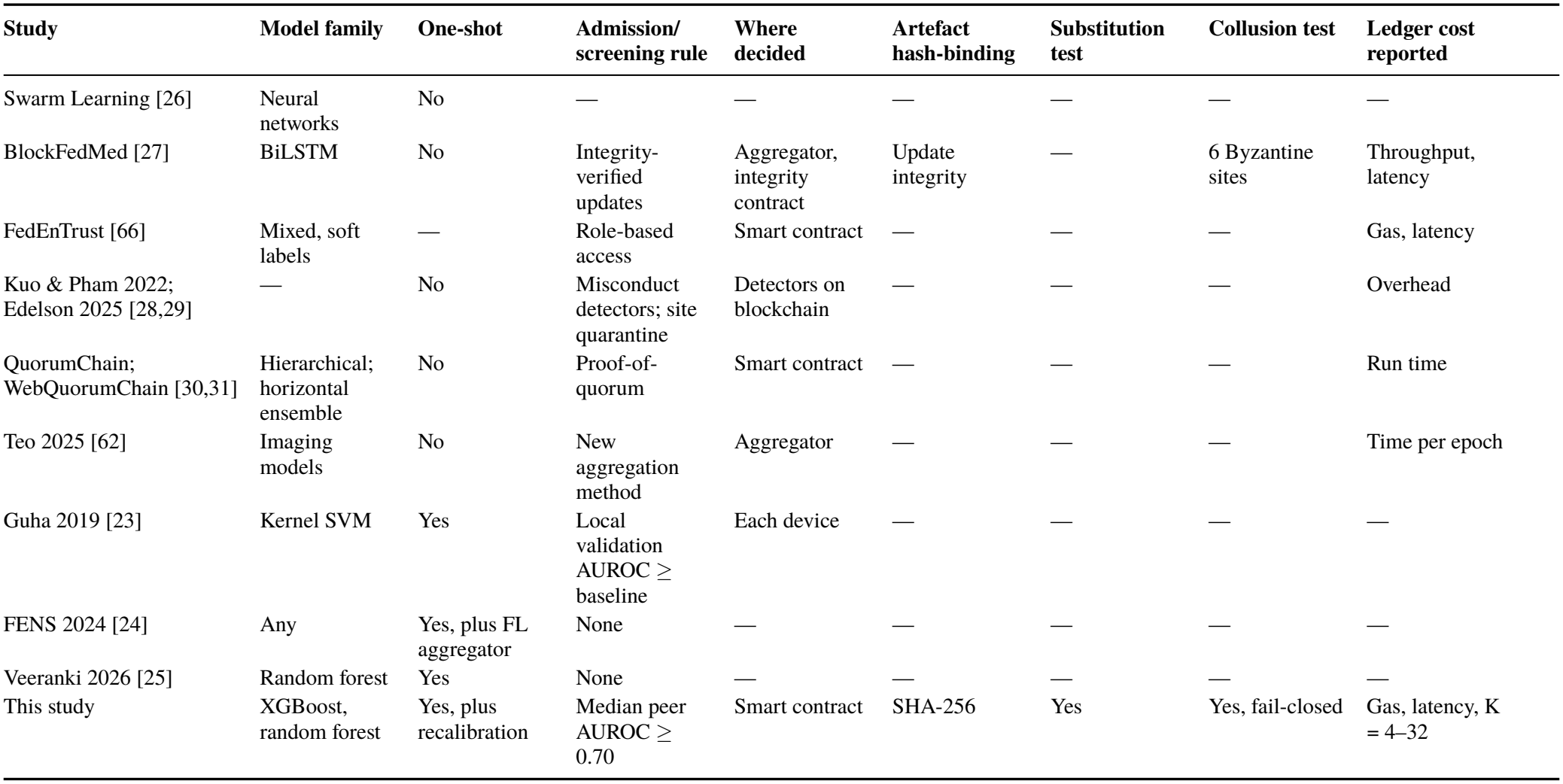
Closest prior designs and this study, from each source’s abstract or text (“—”, not reported).

## 3. Materials and Methods

### 3.1 Data source and cohort

We used the hospital (*hosp*) module of MIMIC-IV v3.1 [32,33,70], de-identified records of patients admitted to Beth Israel Deaconess Medical Center (Boston, MA) in 2008– 2022, accessed through PhysioNet under its credentialed data-use agreement; no further institutional review was required. We included admissions of adults ( ≥18 years) who stayed at least 24 h and were alive at 24 h, so that every prediction is made at a 24 h landmark. The outcome was in-hospital death. Of 546,028 admissions, 120,812 were excluded for stays shorter than 24 h and one for death within 24 h (Supplementary Fig. S1). The cohort comprised **425**,**215 admissions of 186,103 patients**, with 9,984 in-hospital deaths (2.35%); patients contributed a mean of 2.3 admissions.

#### Predictors

Predictors were restricted to information available at 24 h (Supplementary Table S1; 148 columns after one-hot encoding with a consortium-agreed vocabulary): demographics and admission context, utilisation history, twelve Charlson-type comorbidities [71] from *previous* admissions only (index-admission diagnoses are assigned after discharge), and the minimum, maximum and mean of 29 laboratory tests from 6 h before to 24 h after admission. Missingness was treated as informative. Two extended predictor sets were used in a sensitivity analysis (Supplementary Methods S-M1).

### 3.2 Partitions into hospitals

#### Simulated hospitals (primary)

Patients (with all their admissions) were partitioned into four virtual hospitals with three forms of skew:

- **Covariate skew**. *Hospital A*, a geriatric referral centre, receives patients aged ≥70 years at first eligible admission.
- **Case-mix skew**. *Hospital B*, a renal centre, receives the remaining patients with kidney disease (creatinine ≥2.0 mg/dL in any admission, or a chronic kidney disease diagnosis).
- **Feature-availability skew**. The remaining patients are allocated 40:60 to *Hospital C*, a community hospital without lactate, blood-gas, troponin, albumin, liver, bilirubin, differential-count or coagulation panels and with 30% of other results deleted at random, and *Hospital D*, a general hospital.

Each patient follows the rule with probability *p* and is otherwise assigned at random, so *p* = 0 is an IID partition. The primary analysis used *p* = 0.85; sensitivity analyses used *p* = 0 (five seeds) and *p* ∈ {0.5, 1.0} (three seeds). Within each hospital, patients were split 70/10/20 into training, validation and test sets; no patient appeared in more than one hospital or split. Hospital C’s missingness applied to every method, including centralised baselines.

#### Care-era partition (natural)

Each “hospital” was one MIMIC-IV anchor-year group (A: 2008–2010; B: 2011– 2013; C: 2014–2016; D: 2017–2022). The eras differ in case mix, mortality (1.9% to 3.6%) and diagnosis coding (ICD-9 to ICD-10 in 2015); no missingness was added (five seeds).

#### Data-scarce regime

Simulated hospitals hold 71,000– 131,000 admissions, enough for a strong local model. We therefore also kept a random 5%, 10% or 20% of each hos-pital’s training and validation patients (test sets unchanged; five seeds per level), leaving roughly 2,600–4,500 training admissions per hospital at 5%.

### 3.3 Local ensemble members

Each hospital trains two members on its own data: XGBoost [72](early stopping on the hospital’s validation AUPRC) and a random forest [73], with hyper-parameters fixed in advance (Supplementary Methods S-M4). Each serialised member *m* is identified by its digest *h*_*m*_ = SHA-256(bytes_*m*_).

### 3.4 Ledger and admission rule

The audit layer is a Solidity contract, `FederatedAudit-Registry`, written for a permissioned Ethereum consortium and run here on a local single-process Ethereum Virtual Machine [74] (eth-tester), without consensus or network latency. A deployment would use a permissioned EVM network (for example Hyperledger Besu with Quorum Byzantine Fault Tolerance (QBFT) consensus) with one validator per hospital; such consensus tolerates *f* faulty validators when *n* ≥ 3*f* + 1, so four hospitals tolerate one, matching the gate’s tolerance of one false score, but it protects the record, not the truth of a score. A **governor** (the data-governance board) registers hospital nodes; an **aggregator** opens and finalises rounds and commits the global model; **hospital nodes** submit models and attest to peers’ models. Only digests, sample counts and integer scores are stored on-chain. Fig. 1 summarises the data flow, roles and trust boundaries, and Supplementary Fig. S2 the software architecture.

**Fig. 1.**
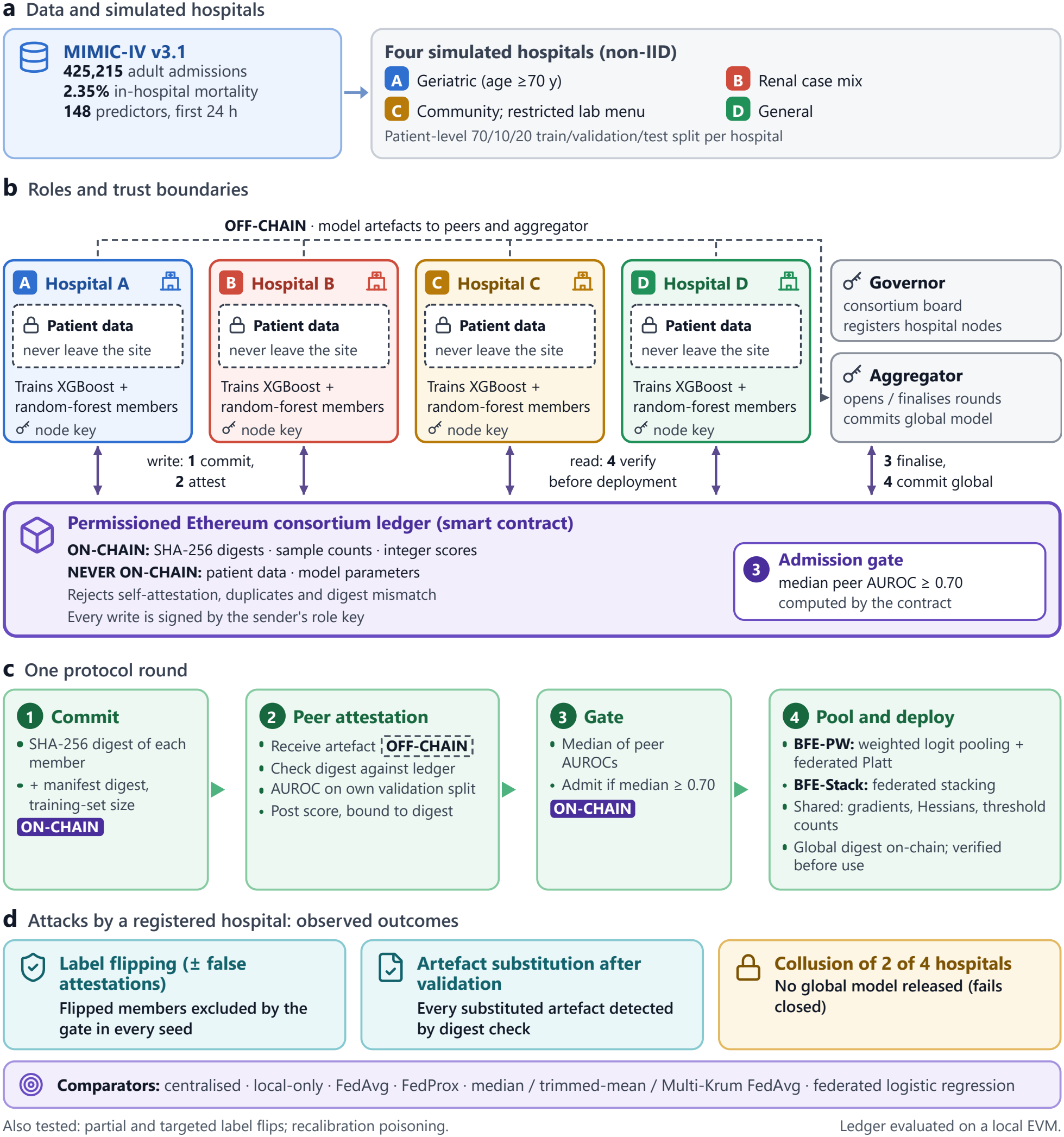
Overview of the Blockchain-Enabled Federated Ensemble (BFE). (a) MIMIC-IV cohort split into four simulated hospitals. (b) Roles and trust boundaries: patient data never leave a hospital, model artefacts are exchanged off-chain, and the consortium ledger stores only digests, sample counts and scores. (c) One round: commit, peer attestation, median-AUROC admission gate, pooling with federated recalibration. (d) Attacks tested and the comparators.

A round proceeds in four steps (functions in Supplementary Methods S-M2). (i) Each hospital commits the digest of each member, a digest of its training manifest (in this proto-type the model family and hospital; a deployment would add the feature schema and hyper-parameters) and its training-set size. The contract accepts one submission per address per round, so each aggregation opens one round per model family; with four hospitals and eight members this takes 37 transactions. (ii) Each peer obtains the artefact off-chain, for example from a content-addressed store, checks its digest, evaluates it on its own validation split and posts the AUROC on-chain; the contract rejects the attestation if the digest the validator reports differs from the committed one, if the validator is the owner, or if it has already attested that model.

(iii) At finalisation the contract computes the **median** peer AUROC *S*_*m*_ of every member and admits the member if it has at least one attestation and *S*_*m*_ ≥ τ. (iv) The aggregator commits the digest of the global specification. A view function `verifyModel` lets the aggregator and each hospital check that a delivered artefact matches an admitted commitment. In the simulation peers scored members in memory, so the delivery check was exercised through verifyModel (attack A3).

The threshold τ = 0.70, a conventional lower bound for acceptable discrimination [75], was fixed before the experiments. When all *K* − 1 = 3 peer scores are present, their median tolerates one false score per model under an honest majority; the contract does not enforce such a quorum, but every model received all three attestations before finalisation. τ is set by the aggregator when it opens a round and is recorded on-chain; it was 0.70 in every round. The contract cannot verify that a validator actually ran the artefact or that the reported AUROC is correct; scores are signed, non-repudiable self-reports. `commitGlobal` does not check the specification against the admission decisions; the admitted set and weights are checked off-chain against the ledger, but recalibration and stacking coefficients only by refitting on validation data.

### 3.5 Aggregation, recalibration and threshold

#### Performance-weighted pooling (BFE-PW, planned as primary)

With *A* the set of admitted members, member weights are

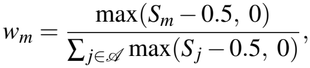

which depend only on on-chain data and can be recom-puted by any participant. Members are pooled on the log-odds scale, *z*(*x*) = ∑_*m*_∈ _*A*_ *w*_*m*_ logit *p*_*m*_(*x*). Because in the clean runs all admitted members had median peer AUROCs between 0.84 and 0.92, the weights were close to uniform (0.11–0.14 for eight members); their practical role is exclusion (zero weight) rather than fine weighting.

Because members are trained under different prevalences (1.4% to 3.8%), the pooled score is recalibrated with a Platt model [76],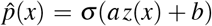, fitted by federated Newton–Raphson in which hospitals share only gradients and Hessians. **BFE-Stack** instead fits an L2-regularised logistic meta-learner over the admitted members’ logits in the same way. A single digest of the global specification (admitted members by hospital and model family, weights and coefficients) is committed on-chain.

#### Recalibration variants

Two further variants were evaluated. In *robust recalibration*, every hospital fits its own Platt pair (*a*_*k*_, *b*_*k*_) on its validation split and shares only those two numbers; the consortium uses their coordinate-wise median, so no single hospital can move the calibration. In the *site-specific intercept*, each hospital keeps the global model and shifts its intercept,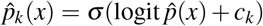, with *c*_*k*_ fitted on its own validation split; nothing is shared and the global model is unchanged.

#### Decision threshold

One F1-maximising threshold per model was chosen from the hospitals’ summed (TP, FP, FN) counts on a 400-point grid; the robust variant used the median of the hospitals’ own thresholds and the site-specific variant each hospital’s own.

### 3.6 Comparators

- **Centralised XGBoost, random forest and multilayer perceptron (MLP)**, trained on the pooled training data (a privacy-violating upper bound). Centralised XGBoost also bounds the accuracy of lossless federated gradient boosting [22].
- **Local-only XGBoost**: each hospital applies its own model to its own patients.
- **FedAvg-MLP** [1] and **FedProx-MLP** (µ = 0.01) [17]: a two-hidden-layer perceptron trained for 40 rounds on federatedly standardised inputs with missingness indicators (Supplementary Methods S-M4).
- **Byzantine-robust FedAvg**: coordinate-wise median (FedMedian) [14] or Multi-Krum with one assumed attacker (FedKrum) [13]. With four hospitals the median equals the trimmed mean that drops one value at each end, so a separate trimmed-mean run (FedTrim) applies the same rule. Multi-Krum’s guarantee needs at least 2*f* + 3 = 5 clients, so with four it is a heuristic.
- **Federated logistic regression (FedLR)**, fitted by federated Newton–Raphson.
- **Uniform ensemble**: the unweighted probability average of all eight tree members, without gate or recalibration.
- **Oracle ensemble** (attack scenarios): BFE with the attacker removed by hand, i.e. a perfect gate.
- **BFE with MLP members** (ablation), separating the aggregation rule from the model family.

### 3.7 Threat model and attack scenarios

The adversary is a registered hospital (D unless stated) that wants to degrade the shared model, globally or for a sub-group. Honest hospitals evaluate faithfully; the aggregator, governor and ledger are trusted in the experiments (a deviating aggregator is discussed in Section 5.2 but not tested), and privacy attacks are out of scope (Section 5.5).

- **A1, label flipping**. D trains its members on inverted out-come labels (under FedAvg, in every local round).
- **A2, false attestation**. As A1, and D also attests an AU-ROC of 0 for every peer model.
- **A3, artefact substitution**. D passes peer validation with an honest model but delivers a label-flipped model at aggregation.
- **A4, collusion**. Hospitals C and D both poison and smear their peers, breaking the honest-majority assumption; included to characterise the failure mode.
- **A5, partial label flipping**. D flips a random 20% or 40% of its training and validation labels, so its models stay informative and may pass the gate.
- **A6, targeted label flipping**. D relabels every death among its female patients as a survival, aiming to suppress risk for one subgroup while keeping overall discrimination.
- **A7, recalibration and threshold poisoning**. D’s models are honest, but it computes its contributions to the federated Platt fit, the stacking fit and the threshold counts from inverted validation labels. These channels bypass the admission gate.

A tamper test applied 200 single-bit flips, 200 one-byte appends and 20 retrained substitutes to a small model.

### 3.8 Evaluation and statistical analysis

Discrimination was measured by AUROC and, given the 2.35% outcome rate, by AUPRC [77]. Calibration was assessed by the calibration intercept (calibration-in-the-large), calibration slope and integrated calibration index (ICI; mean absolute difference between predicted risk and lowess-smoothed observed risk) [78], pooled and per hospital, by the observed-to-expected (O/E) ratio, by the Brier score [41] and by the expected calibration error (ECE). Sensitivity and positive predictive value at the federated threshold, and decision curves [79], describe clinical use.

A seed re-draws the partition, splits and initialisation, so the SD across seeds reflects partition and training variability. **Primary inference** was a joint patient-clustered bootstrap: in each of 1,000 replicates, patients (with all their admissions) were resampled in each seed’s test set, the between-method difference was averaged over the five seeds, and the 95% percentile interval of that mean was reported. Planned comparisons were BFE-PW and BFE-Stack against FedAvg, FedProx, FedMedian, local-only and centralised XGBoost, for AUROC and AUPRC. As a secondary analysis, DeLong’s test [80] compared BFE-PW per seed with FedAvg, FedProx, local-only and centralised XGBoost, the uniform ensemble and BFE-Stack, with Holm adjustment [81] over the 30 tests. A subgroup audit covered sex, age band and race group (strata with ≥ 20 deaths). Ledger cost was measured as gas and latency per transaction for 4–32 hospitals. Software is listed in Supplementary Methods S-M3; reporting follows TRIPOD+AI [40].

### 3.9 Use of AI tools

The study used an AI coding assistant (an agentic large-language-model tool) from September to November 2026. Under the first author’s instructions it wrote and debugged the analysis code, ran the analyses on the authors’ workstation, checked references against PubMed and Crossref, and drafted and revised the text. Patient-level data stayed on the workstation (PhysioNet data use agreement); the assistant received code, aggregate results and log summaries only. Every number was produced by the authors’ scripts and checked against the result files, and the authors take responsibility for all content. The instructions given to the assistant are summarised in Supplementary File S2.

## 4. Results

### 4.1 Cohort and induced heterogeneity

In the primary configuration (*p* = 0.85) the four simulated hospitals differed in size, case mix, mortality (1.36% at D to 3.83% at A), median age (53 years at C and D to 80 at A) and missing laboratory features (36.6% at B to 73.7% at C) (Table 2, Fig. 2 a–c); under the IID partition mortality ranged only from 2.32% to 2.38% (Supplementary Table S2). The pooled test set held on average 85,227 admissions and 1,956 deaths per seed.

**Table 2.** Characteristics of the four simulated hospitals (purity *p* = 0.85, seed 0). IQR, interquartile range; CKD, chronic kidney disease.

| **Hospital** | **Admissions** | **Patients** | **Deaths, n (%)** | **Age, median (IQR)** | **Female, %** | **Emergency admission, %** | **Creatinine max ≥2.0 mg/dL, %** | **Prior CKD, %** | **Lab features missing, %** |
|---|---|---|---|---|---|---|---|---|---|
| Hospital A (geriatric referral) | 130,053 | 57,740 | 4,977 (3.83) | 80 (74–86) | 53.3 | 48.1 | 11.7 | 20.4 | 39.9 |
| Hospital B (renal/ nephrology) | 71,426 | 17,011 | 1,908 (2.67) | 61 (52–68) | 43.3 | 49.0 | 31.8 | 45.8 | 36.6 |
| Hospital C (community, sparse labs) | 92,417 | 46,032 | 1,313 (1.42) | 53 (38–63) | 55.0 | 38.9 | 1.7 | 2.7 | 73.7 |
| Hospital D (general acute care) | 131,319 | 65,320 | 1,786 (1.36) | 53 (37–63) | 55.8 | 38.5 | 1.2 | 1.8 | 51.2 |
| Pooled | 425,215 | 186,103 | 9,984 (2.35) | 63 (48–75) | 52.8 | 43.3 | 9.6 | 15.1 | 50.2 |

**Fig. 2.**
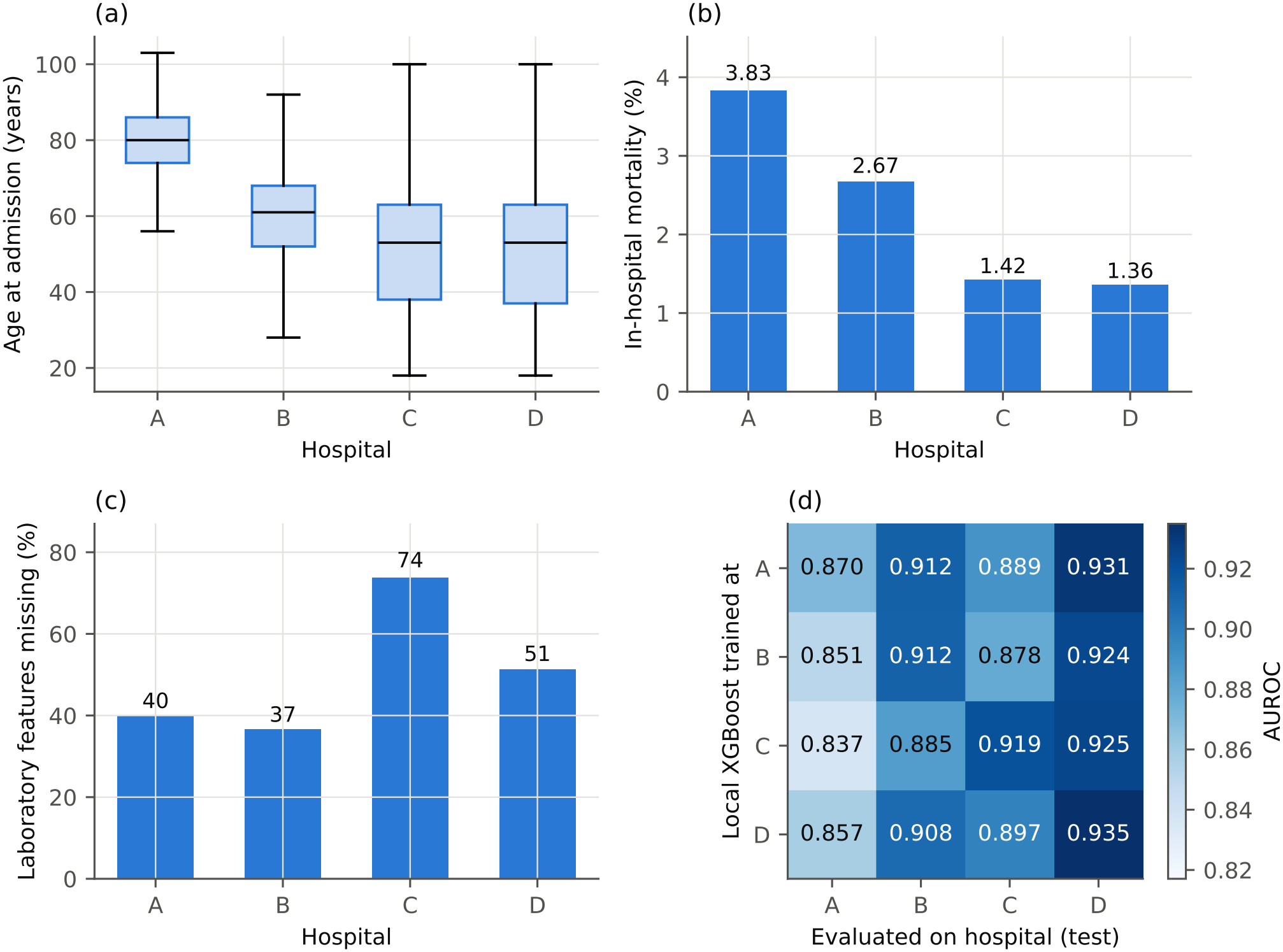
Induced heterogeneity at *p* = 0.85 (seed 0): (a) age, (b) in-hospital mortality and (c) missing laboratory features by hospital; (d) AUROC of each hospital’s local XGBoost model (rows) on each hospital’s test patients (columns), mean of five seeds.

The partition induced real distribution shift (Fig. 2 d): on Hospital C’s sparsely measured patients, other sites’ models reached AUROCs of 0.878–0.897 against 0.919 for C’s own model.

### 4.2 Performance with large simulated hospitals

Table 3 summarises performance at *p* = 0.85 and Table 4 the primary inference. Centralised XGBoost reached the highest discrimination (AUROC 0.915, AUPRC 0.337), and with 71,000–131,000 admissions per hospital local-only XG-Boost was already strong (0.910). In the joint patient-clustered bootstrap, BFE-PW’s AUROC did not differ from FedAvg’s (Δ +0.0001, 95% CI −0.0014 to 0.0018) and was lower than FedProx’s, local-only XGBoost’s and cen-tralised XGBoost’s; BFE-Stack was higher than FedAvg (Δ +0.0055, 95% CI 0.0042 to 0.0070), similar to FedProx and lower than centralised XGBoost. Both ensembles had lower AUPRC than FedAvg and FedProx (BFE-PW 0.301, BFE-Stack 0.309, FedAvg 0.315, FedProx 0.322; Table 4). The secondary per-seed DeLong analysis agreed (Supplementary Table S3).

**Table 3.** Pooled-test performance with large simulated hospitals (*p* = 0.85; mean ± SD over five seeds; calibration measures are five-seed means). *Centralised models pool all records and are an upper reference. ICI, integrated calibration index. Sensitivity at the federated F1-optimal threshold.

| **Method** | **AUROC** | **AUPRC** | **Sensitivity** | **Brier** | **Calibration intercept** | **Calibration slope** | **ICI** |
|---|---|---|---|---|---|---|---|
| Centralised XGBoost* | 0.915 ± 0.003 | 0.337 ± 0.012 | 0.405 ± 0.052 | 0.0181 ± 0.0005 | 0.02 | 0.99 | 0.0022 |
| Centralised random forest* | 0.897 ± 0.004 | 0.295 ± 0.009 | 0.384 ± 0.034 | 0.0189 ± 0.0006 | – | – | – |
| Centralised MLP* | 0.905 ± 0.002 | 0.319 ± 0.012 | 0.362 ± 0.029 | 0.0184 ± 0.0005 | – | – | – |
| Local-only XGBoost | 0.910 ± 0.004 | 0.315 ± 0.011 | 0.384 ± 0.038 | 0.0185 ± 0.0006 | 0.03 | 1.01 | 0.0026 |
| FedAvg | 0.902 ± 0.003 | 0.315 ± 0.012 | 0.360 ± 0.055 | 0.0186 ± 0.0005 | −0.09 | 0.88 | 0.0033 |
| FedProx | 0.906 ± 0.003 | 0.322 ± 0.011 | 0.382 ± 0.039 | 0.0184 ± 0.0005 | −0.10 | 0.93 | 0.0026 |
| Uniform ensemble | 0.900 ± 0.004 | 0.303 ± 0.010 | 0.358 ± 0.031 | 0.0188 ± 0.0006 | – | – | – |
| BFE with MLP members (ablation) | 0.894 ± 0.004 | 0.298 ± 0.007 | 0.345 ± 0.034 | 0.0188 ± 0.0006 | – | – | – |
| BFE-Stack | 0.907 ± 0.004 | 0.309 ± 0.008 | 0.364 ± 0.033 | 0.0186 ± 0.0005 | −0.03 | 1.01 | 0.0023 |
| BFE-PW | 0.902 ± 0.004 | 0.301 ± 0.010 | 0.341 ± 0.033 | 0.0187 ± 0.0005 | −0.03 | 1.01 | 0.0024 |

**Table 4.** Primary inference: differences in AUROC and AUPRC (first method minus second) with 95% CIs from 1,000 joint patient-clustered bootstrap replicates over the five seeds’ test sets, and the number of seeds in which the difference was positive.

| **Comparison** | **ΔAUROC (95% CI)** | **ΔAUROC > 0, seeds** | **ΔAUPRC (95% CI)** | **ΔAUPRC > 0, seeds** |
|---|---|---|---|---|
| BFE-PW − FedAvg-MLP | +0.0001 (−0.0014 to +0.0018) | 3/5 | −0.0136 (−0.0181 to −0.0090) | 0/5 |
| BFE-PW − FedProx-MLP | −0.0047 (−0.0060 to −0.0033) | 0/5 | −0.0214 (−0.0253 to −0.0175) | 0/5 |
| BFE-PW − Local-XGB | −0.0083 (−0.0096 to −0.0069) | 0/5 | −0.0141 (−0.0183 to −0.0096) | 0/5 |
| BFE-PW − Central-XGB | −0.0128 (−0.0140 to −0.0114) | 0/5 | −0.0356 (−0.0398 to −0.0310) | 0/5 |
| BFE-PW − FedMedian-MLP | +0.0004 (−0.0011 to +0.0020) | 4/5 | −0.0131 (−0.0177 to −0.0088) | 0/5 |
| BFE-Stack − FedAvg-MLP | +0.0055 (+0.0042 to +0.0070) | 5/5 | −0.0052 (−0.0101 to −0.0006) | 2/5 |
| BFE-Stack − FedProx-MLP | +0.0007 (−0.0005 to +0.0019) | 3/5 | −0.0130 (−0.0171 to −0.0087) | 0/5 |
| BFE-Stack − Local-XGB | −0.0029 (−0.0041 to −0.0018) | 0/5 | −0.0057 (−0.0099 to −0.0012) | 1/5 |
| BFE-Stack − Central-XGB | −0.0074 (−0.0084 to −0.0063) | 0/5 | −0.0272 (−0.0312 to −0.0228) | 0/5 |
| BFE-Stack − FedMedian-MLP | +0.0058 (+0.0044 to +0.0073) | 5/5 | −0.0047 (−0.0093 to −0.0001) | 2/5 |

With MLP members, BFE reached 0.894 against 0.902 for FedAvg with the same architecture, so one-round ensembling cost about 0.008 AUROC and tree members recovered it.

Pooled calibration was good for all federated methods (BFE-PW intercept −0.03, slope 1.01, ICI 0.002; Fig. 3 c), but it hid site-level errors. BFE-PW over-predicted deaths at Hospital B (O/E 0.64) and under-predicted them at Hospital C by about half (O/E 2.13); BFE-Stack (0.72 and 1.82), FedAvg (0.74 and 1.71) and FedProx (0.74 and 1.57) erred in the same direction, while centralised and local-only XG-Boost stayed within 0.89–1.09 at every hospital (Supplementary Table S4). With one consortium-wide threshold, BFE-PW therefore identified only 3.3% of deaths at Hospital C, against 27.5% for centralised XGBoost. Section 4.5 tests two corrections.

**Fig. 3.**
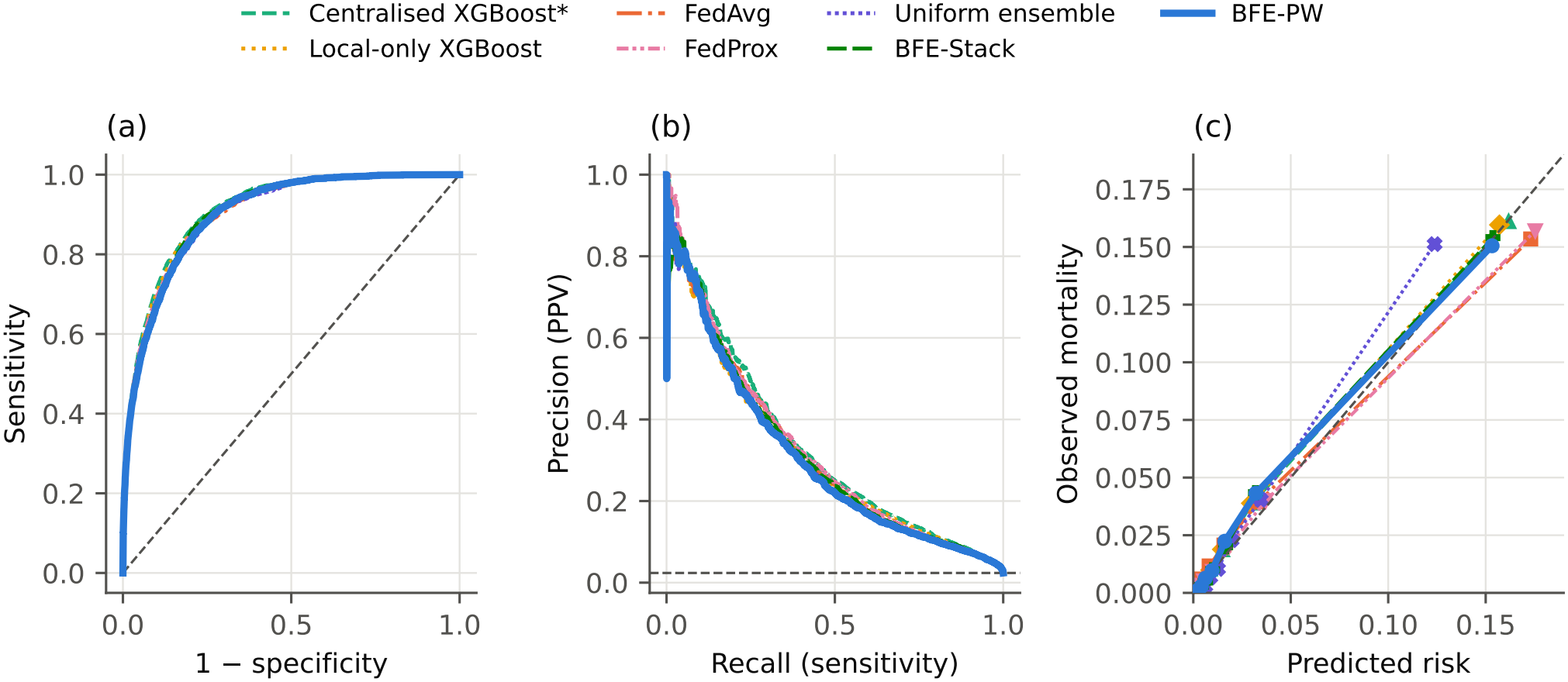
Pooled-test (a) ROC curves, (b) precision–recall curves and (c) reliability diagrams (deciles of predicted risk), seed 0, *p* = 0.85. Five-seed means are given in Table 3.

Per hospital (Table 5), BFE-PW matched each hospital’s own model within 0.004 AUROC; at Hospital C, the ensembles and local XGBoost outperformed FedAvg (0.918 against 0.896), consistent with trees tolerating structurally missing panels.

**Table 5.** Per-hospital AUROC and AUPRC (*p* = 0.85; mean ± SD over five seeds). Local-only XGBoost applies each hospital’s model to its own patients.

| **Method** | **A AUROC** | **A AUPRC** | **B AUROC** | **B AUPRC** | **C AUROC** | **C AUPRC** | **D AUROC** | **D AUPRC** |
|---|---|---|---|---|---|---|---|---|
| Local-only XGBoost | 0.870 ± 0.005 | 0.323 ± 0.011 | 0.912 ± 0.008 | 0.401 ± 0.019 | 0.919 ± 0.003 | 0.223 ± 0.018 | 0.935 ± 0.007 | 0.271 ± 0.029 |
| FedAvg | 0.864 ± 0.004 | 0.322 ± 0.014 | 0.910 ± 0.006 | 0.414 ± 0.024 | 0.896 ± 0.006 | 0.180 ± 0.013 | 0.933 ± 0.007 | 0.309 ± 0.031 |
| FedProx | 0.868 ± 0.005 | 0.330 ± 0.014 | 0.914 ± 0.008 | 0.418 ± 0.025 | 0.907 ± 0.006 | 0.203 ± 0.023 | 0.936 ± 0.007 | 0.312 ± 0.035 |
| Uniform ensemble | 0.865 ± 0.003 | 0.316 ± 0.011 | 0.909 ± 0.008 | 0.404 ± 0.009 | 0.915 ± 0.005 | 0.236 ± 0.031 | 0.931 ± 0.009 | 0.297 ± 0.032 |
| BFE-PW | 0.866 ± 0.004 | 0.315 ± 0.011 | 0.912 ± 0.008 | 0.406 ± 0.009 | 0.918 ± 0.003 | 0.227 ± 0.027 | 0.935 ± 0.008 | 0.298 ± 0.031 |
| Centralised XGBoost* | 0.875 ± 0.003 | 0.332 ± 0.013 | 0.915 ± 0.006 | 0.425 ± 0.012 | 0.924 ± 0.001 | 0.243 ± 0.025 | 0.941 ± 0.006 | 0.326 ± 0.036 |

### 4.3 Data-scarce hospitals and natural care-era sites

When each hospital kept only part of its patients, collaboration became useful (Table 6, Fig. 4). With 5% of the data (about 3,700 training admissions per hospital), local-only XGBoost fell to an AUROC of 0.844 ± 0.026, whereas BFE-PW reached 0.866 ± 0.005 and BFE-Stack 0.866 ± 0.007, similar to FedAvg (0.863) and FedProx (0.867); BFE-PW exceeded the local models in all five seeds (by 0.005–0.058). At 10% (about 7,400 admissions) the federated methods were 0.000–0.004 above local models, and at 20% (about 14,800 admissions) local models had caught up (0.890 against 0.887 for BFE-PW and 0.892 for FedProx). The gate admitted all eight honest members in every seed and regime. In AUPRC the ensembles stayed below FedAvg and FedProx in every data-scarce regime and with large simulated hospitals.

**Table 6.** AUROC (mean ± SD over five seeds) and, in parentheses, mean AUPRC by training-data regime and partition. Percentages are the share of each simulated hospital’s training and validation patients kept (test sets unchanged); care-era sites are MIMIC-IV anchor-year groups. *Upper reference.

| **Method** | **5% (3,704 per site)** | **10% (7,399 per site)** | **20% (14,825 per site)** | **100% (74,460 per site)** | **Care-era sites** |
|---|---|---|---|---|---|
| Local-only XGBoost | 0.844 ± 0.026 (0.194) | 0.876 ± 0.008 (0.239) | 0.890 ± 0.007 (0.269) | 0.910 ± 0.004 (0.315) | 0.911 ± 0.003 (0.322) |
| Centralised XGBoost* | 0.883 ± 0.010 (0.250) | 0.897 ± 0.004 (0.281) | 0.904 ± 0.003 (0.305) | 0.915 ± 0.003 (0.337) | 0.918 ± 0.003 (0.351) |
| FedAvg | 0.863 ± 0.007 (0.231) | 0.876 ± 0.007 (0.254) | 0.886 ± 0.005 (0.275) | 0.902 ± 0.003 (0.315) | 0.912 ± 0.002 (0.336) |
| FedProx | 0.867 ± 0.007 (0.238) | 0.880 ± 0.006 (0.263) | 0.892 ± 0.004 (0.292) | 0.906 ± 0.003 (0.322) | 0.914 ± 0.003 (0.341) |
| BFE-PW | 0.866 ± 0.005 (0.222) | 0.878 ± 0.005 (0.241) | 0.887 ± 0.005 (0.265) | 0.902 ± 0.004 (0.301) | 0.913 ± 0.004 (0.326) |
| BFE-Stack | 0.866 ± 0.007 (0.218) | 0.880 ± 0.003 (0.246) | 0.889 ± 0.005 (0.270) | 0.907 ± 0.004 (0.309) | 0.916 ± 0.003 (0.338) |

**Fig. 4.**
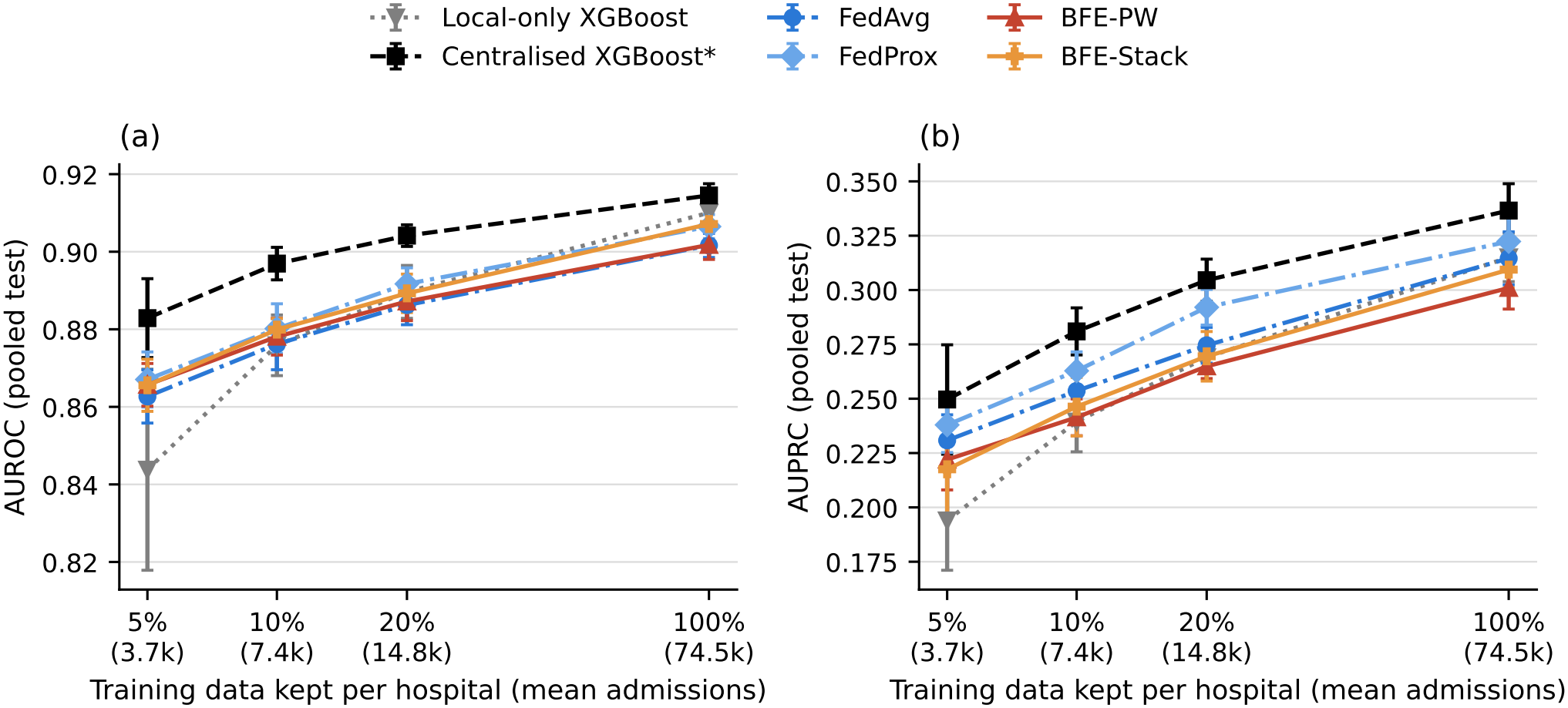
Pooled-test (a) AUROC and (b) AUPRC against the amount of training data kept per hospital (mean ± SD over five seeds; log axis; the 100% point is the main configuration). *Upper reference.

On the care-era partition, where sites differ by calendar period, case mix and diagnosis coding rather than by a rule, BFE-Stack reached an AUROC of 0.916 and BFE-PW 0.913, against 0.912 for FedAvg, 0.914 for FedProx, 0.911 for local-only and 0.918 for centralised XGBoost (Table 6). In the joint bootstrap (Supplementary Table S5), BFE-Stack exceeded FedAvg, FedProx and local-only XGBoost (by 0.0043, 0.0021 and 0.0050; all 95% CIs above zero) and was below centralised XGBoost; BFE-PW did not differ from FedAvg or local-only models. Local-only models no longer outperformed federation.

### 4.4 Poisoning, substitution and collusion

Table 7 and Fig. 5 report the attacks, with Hospital D as the adversary.

**Table 7.** Attack scenarios with Hospital D as adversary (mean ± SD over five seeds). A1–A4 rows come from the main runs; A5–A7, the oracle and the robust-aggregation rows from separate runs on the same partitions and splits. The oracle excludes D by hand. With four hospitals, trimmed-mean FedAvg applies the same rule as the coordinate-wise median FedAvg quoted in the text (a separate run).

| **Scenario** | **Method** | **AUROC** | **AUPRC** | **Sensitivity** | **Note** |
|---|---|---|---|---|---|
| Clean | BFE-PW | 0.902 ± 0.004 | 0.301 ± 0.010 | 0.341 ± 0.033 |  |
| Clean | FedAvg | 0.902 ± 0.003 | 0.315 ± 0.012 | 0.360 ± 0.055 |  |
| A1 label flip | BFE-PW | 0.902 ± 0.004 | 0.301 ± 0.010 | 0.344 ± 0.036 |  |
| A1 label flip | FedAvg | 0.868 ± 0.011 | 0.224 ± 0.033 | 0.322 ± 0.012 |  |
| A1 label flip | Uniform ensemble (no gate) | 0.885 ± 0.005 | 0.286 ± 0.009 | 0.332 ± 0.039 |  |
| A2 flip + false attestation | BFE-PW | 0.902 ± 0.004 | 0.301 ± 0.010 | 0.356 ± 0.041 |  |
| A3 artefact substitution | BFE-PW (digest check) | 0.902 ± 0.004 | 0.301 ± 0.010 | 0.344 ± 0.036 |  |
| A3 artefact substitution | Same, no digest check | 0.895 ± 0.004 | 0.287 ± 0.011 | 0.332 ± 0.035 |  |
| A4 collusion (C and D) | Uniform ensemble (no gate) | 0.491 ± 0.012 | 0.158 ± 0.015 | 0.215 ± 0.013 | BFE released no model |
| Clean | Federated LR | 0.895 ± 0.003 | 0.272 ± 0.008 | 0.389 ± 0.028 |  |
| Clean | Trimmed-mean FedAvg | 0.901 ± 0.004 | 0.312 ± 0.011 | 0.374 ± 0.027 |  |
| A1 label flip | Trimmed-mean FedAvg | 0.889 ± 0.005 | 0.285 ± 0.014 | 0.353 ± 0.037 |  |
| Clean | Multi-Krum FedAvg | 0.901 ± 0.003 | 0.314 ± 0.011 | 0.359 ± 0.032 |  |
| A1 label flip | Multi-Krum FedAvg | 0.901 ± 0.003 | 0.311 ± 0.011 | 0.377 ± 0.054 |  |
| Oracle (D removed by hand) | BFE-PW | 0.902 ± 0.004 | 0.301 ± 0.010 | 0.344 ± 0.036 |  |
| A5 partial flip 20% | BFE-PW | 0.902 ± 0.004 | 0.300 ± 0.009 | 0.353 ± 0.042 |  |
| A5 partial flip 20% | FedAvg | 0.892 ± 0.006 | 0.305 ± 0.016 | 0.341 ± 0.041 |  |
| A5 partial flip 40% | BFE-PW | 0.902 ± 0.004 | 0.300 ± 0.009 | 0.349 ± 0.040 |  |
| A5 partial flip 40% | FedAvg | 0.861 ± 0.013 | 0.269 ± 0.010 | 0.290 ± 0.016 |  |
| A6 targeted flip (female deaths) | BFE-PW | 0.896 ± 0.005 | 0.282 ± 0.010 | 0.363 ± 0.032 |  |
| A6 targeted flip (female deaths) | FedAvg | 0.894 ± 0.003 | 0.294 ± 0.013 | 0.381 ± 0.020 |  |
| A7 recalibration poisoning | BFE-PW (federated Platt) | 0.098 ± 0.004 | 0.012 ± 0.000 | 0.043 ± 0.007 |  |
| A7 recalibration poisoning | BFE-PW<br>(median-of-sites Platt) | 0.902 ± 0.004 | 0.301 ± 0.010 | 0.465 ± 0.042 |  |
| Clean | BFE-PW<br>(median-of-sites Platt) | 0.902 ± 0.004 | 0.301 ± 0.010 | 0.357 ± 0.031 |  |
| Clean | BFE-PW (site-specific intercept) | 0.908 ± 0.004 | 0.312 ± 0.011 | 0.370 ± 0.023 |  |

**Fig. 5.**
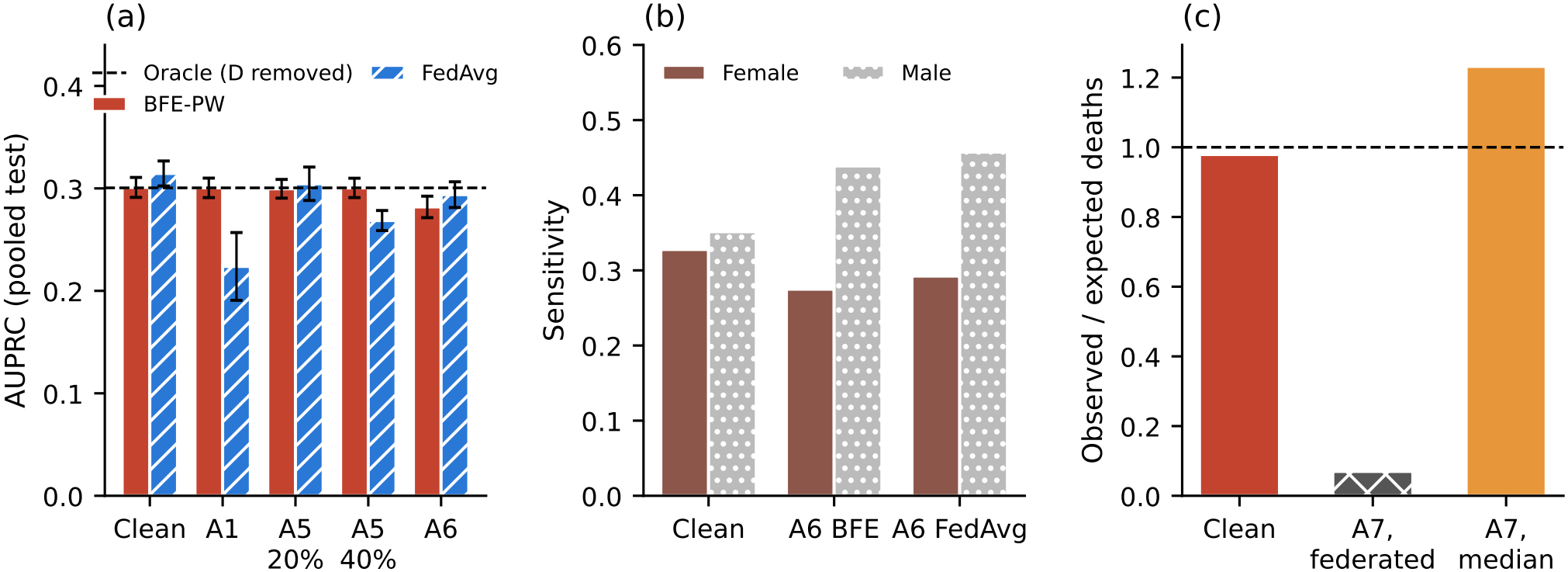
Stealthier attacks (Hospital D adversary; five seeds). (a) AUPRC of BFE-PW and FedAvg under label-flipping attacks; dashed line, oracle ensemble without D. (b) Sensitivity among female and male patients under the targeted attack A6. (c) Pooled observed-to-expected deaths under recalibration poisoning (A7) with federated and median-of-sites Platt recalibration; dashed line, perfect calibration-in-the-large.

#### Gross label flipping (A1, A2)

FedAvg lost 29% of its AUPRC (0.315 to 0.224) and its AUROC fell to 0.868. Coordinate-wise median aggregation protected in part (AU-ROC 0.884, AUPRC 0.274; trimmed mean in a separate run 0.889 and 0.285), whereas Multi-Krum fully protected Fe-dAvg (0.901, 0.311 against 0.901, 0.314 without attack). The ungated uniform ensemble lost some discrimination (AU-ROC 0.900 to 0.885) and became badly miscalibrated (Brier 0.077 against 0.019). Under BFE, peers scored D’s poisoned members at median AUROCs of 0.10–0.13; the contract rejected both members in every seed, admitted all six honest members, and BFE-PW was unchanged (AUROC 0.902, AUPRC 0.301), equal to the oracle ensemble that removes D by hand (0.902 and 0.301). False attestations of 0 by D (A2) lowered honest members’ medians by at most 0.07 and changed no decision.

#### Partial flipping (A5)

When D flipped 20% of its deaths and survivals, its members still scored median peer AUROCs of 0.75–0.82 and were admitted in every seed. They did no measurable harm: BFE-PW (AUROC 0.902, AUPRC 0.300) and BFE-Stack (0.906, 0.307) equalled the oracle ensemble, because the poisoned members kept useful ranking and received lower weights. FedAvg lost 0.009 AUROC (0.892) and its calibration worsened (ECE 0.027 against 0.003). With 40% flipped, D’s members scored 0.57–0.72 and were re-jected in 9 of 10 cases (Supplementary Table S6); BFE was again equal to the oracle, whereas FedAvg fell to 0.861 (AUPRC 0.269).

#### Targeted flipping (A6)

When D relabelled the deaths of its female patients as survivals, its XGBoost member scored median peer AUROCs of 0.71–0.73, just above the threshold, and its random forest 0.82–0.85; both were admitted in every seed. The attack lowered BFE-PW’s pooled AUROC only from 0.902 to 0.896 (AUPRC 0.301 to 0.282) and left its AUROC in women almost unchanged (0.915 to 0.913), but at the shared threshold the sensitivity for deaths in women fell from 32.8% to 27.5% while that in men rose from 35.1% to 43.9% (Supplementary Table S7). FedAvg was affected similarly. BFE-Stack was more resistant (AUROC unchanged at 0.907; sensitivity in women 35.8% to 33.2%). A gate based on pooled discrimination therefore did not detect a targeted attack that shifted harm onto one group.

#### Recalibration poisoning (A7)

When D’s members were honest but D submitted flipped labels to the recalibration and threshold steps, which the gate does not inspect, the pooled Platt fit inverted the score (slope − 0.27 to −0.31 in the five seeds): BFE-PW’s AUROC fell to 0.098 and BFE-Stack’s to 0.252, and BFE-PW flagged 54–60% of patients while finding 4.3% of deaths. Replacing the pooled fit with the median of the four sites’ Platt coefficients removed the effect on discrimination (AUROC 0.902, AUPRC 0.301) and calibration (ECE 0.005), but D could still shift the median-of-sites threshold: sensitivity rose to 46.5% (35.7% without attack) and positive predictive value fell to 23.9% (34.6% for standard BFE-PW without attack).

#### Artefact substitution (A3)

All 10 substituted artefacts (two per seed) failed verifyModel and were excluded; without the digest check, the same ensemble would have deployed them and its AUROC would have fallen to 0.895 (AUPRC 0.287). In a separate tamper test, the ledger detected 200 of 200 single-bit flips, 200 of 200 one-byte appends and 20 of 20 retrained substitutes. These results confirm that the digest comparison works, and follow from SHA-256 collision resistance, not from a blockchain-specific property.

#### Collusion (A4)

With two of four hospitals poisoning and attesting zero for every honest model, no member reached the threshold and the contract released no global model in any seed. An ungoverned uniform ensemble in the same setting fell to an AUROC of 0.491. BFE therefore fails closed when its honest-majority assumption is broken.

### 4.5 Correcting site-level miscalibration

Letting each hospital fit its own intercept on its validation data, with no data shared, removed the site-level bias of BFE-PW (Supplementary Table S4): O/E moved from 2.13 to 0.93 at Hospital C, from 0.64 to 0.98 at Hospital B, and from 1.05 and 0.95 to 0.99 and 1.00 at A and D; the share of deaths identified at Hospital C rose from 3.3% to 35.5%. Per-site AU-ROCs were unchanged, and because the corrected risks are comparable across hospitals the pooled AUROC rose from 0.902 to 0.908 (AUPRC 0.301 to 0.312). The median-of-sites recalibration, which protects against poisoned recalibration (Section 4.4), did not help here: without attack it made Hospital C’s under-prediction worse (O/E 3.12) and shifted pooled calibration (O/E 1.14, slope 0.89). The two correc-tions are complementary: the robust fit for the shared model, the local intercept at each hospital.

### 4.6 Subgroups

Discrimination varied more across age bands than across sex or race for every method (Fig. 6): BFE-PW’s AUROC fell from 0.957 (18–49 years) to 0.832 ( ≥ 85 years), as did centralised XGBoost’s (0.965 to 0.841), and the largest gap between sexes or race groups was 0.032. BFE-PW underpredicted mortality at ≥ 85 years (O/E 1.17) and in patients with unrecorded race (1.28), where FedAvg did not (0.99), and over-predicted it at 18–49 years (0.76) and in Black (0.81) and Hispanic (0.83) patients; BFE-Stack’s O/E stayed within 0.89–1.15 (Supplementary Table S8).

**Fig. 6.**
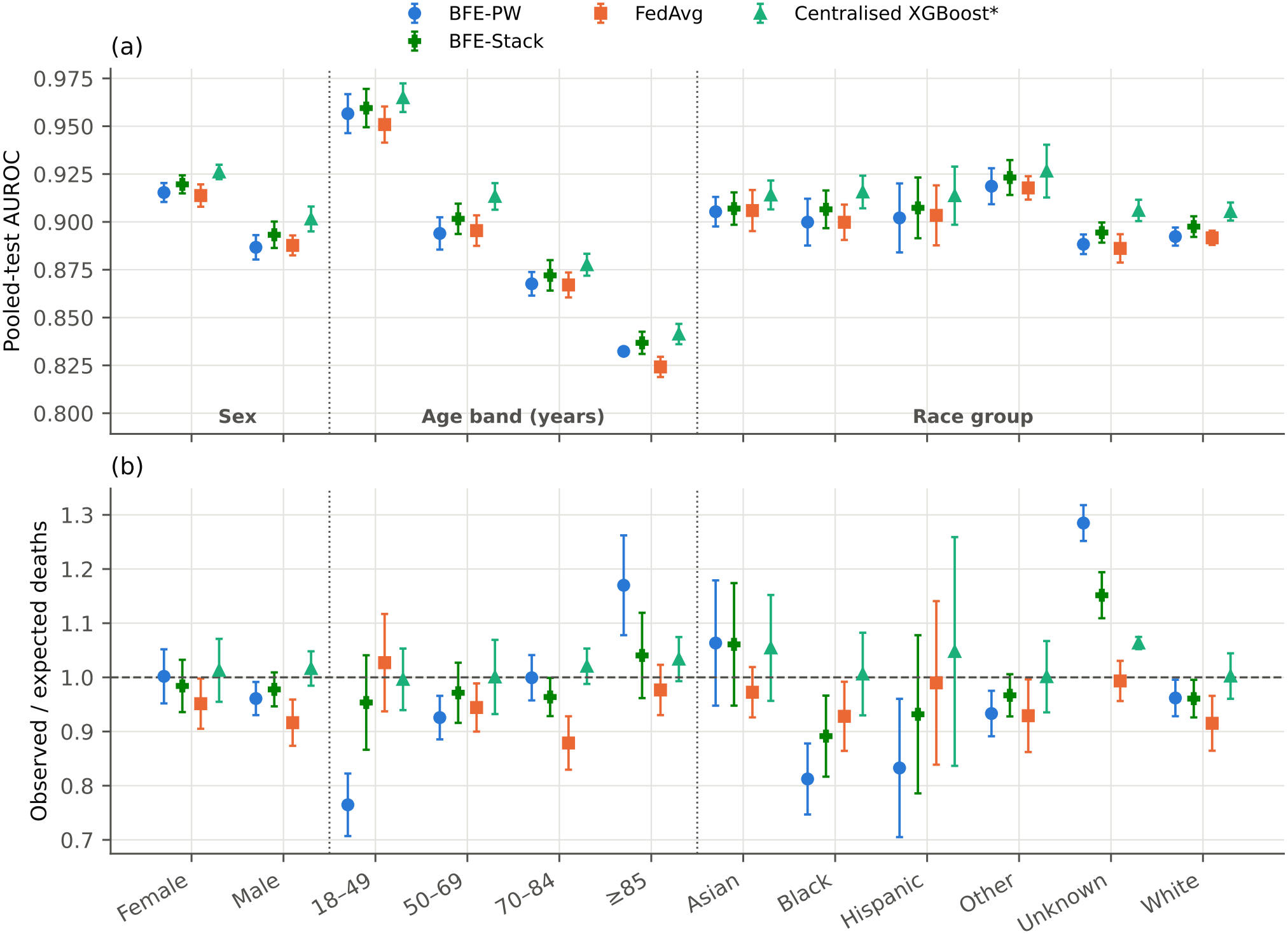
Subgroup audit (*p* = 0.85; mean ± SD over five seeds): (a) AUROC and (b) observed-to-expected deaths by sex, age band and race group.

### 4.7 Ledger cost and scalability

Deploying the contract cost 1,147,766 gas. One aggregation of eight members required 37 transactions and about 3.28 million gas (Supplementary Table S9), with 92–285 ms per transaction on the local EVM. The protocol added about 30 s to local tree training, and artefacts were exchanged once (0.56 MB per XGBoost and 8.9 MB per random-forest member) instead of weights over 40 rounds.

Cost grows with consortium size *K* because attestations scale as *K*(*K*− 1) (Table 8, Supplementary Fig. S3): at *K* = 32 a round needed 1,027 transactions and 77.8 million gas, and one finalizeRound call 4.99 million gas. Because a permissioned network sets its own gas limit, these figures show relative scaling.

**Table 8.** Ledger cost by consortium size for one model family (one model per hospital per round; local Ethereum Virtual Machine). The two-family protocol used in this study roughly doubles each figure.

| **Hospitals (K)** | **Transactions per round** | **Gas per round (millions)** | **finalizeRound gas** | **Round time on local EVM (s)** |
|---|---|---|---|---|
| 4 | 19 | 1.68 | 103,853 | 5.2 |
| 8 | 67 | 5.42 | 238,124 | 20.9 |
| 16 | 259 | 19.75 | 871,202 | 127.6 |
| 32 | 1,027 | 77.76 | 4,987,479 | 502.6 |

### 4.8 Sensitivity analyses

As skew increased from IID to deterministic referral, FedAvg (0.897 to 0.903) and BFE-Stack (0.906–0.907) were stable and BFE-PW fell to 0.896 at *p* = 1 (Supplementary Fig. S4, Supplementary Table S10). Adding ICU, medication and microbiology predictors raised every method’s AUROC by 0.009–0.013, and the extended set plus vital signs by 0.013–0.018 (both relative to the primary predictors), with the same ordering (Supplementary Table S11); they narrowed but did not remove the under-prediction at Hospital C (O/E 1.38 with vital signs). Decision curves showed positive net benefit for all federated models at thresholds of 1–30% (Supplementary Fig. S5).

## 5. Discussion

### 5.1 Principal findings

Four findings stand out. First, the protocol’s accuracy cost was small but real: with large hospitals the ensembles matched FedAvg or FedProx in AUROC but stayed below local-only and centralised XGBoost, and below FedAvg and FedProx in AUPRC. Second, collaboration paid off for small hospitals: at 5% of the data every federated method beat local models by about 0.02 AUROC, whereas above roughly 15,000 training admissions local models were as good. Third, the gate excluded grossly poisoned and sub-stituted models and failed closed under collusion, but attacks that kept validation discrimination high passed it, and two did harm. Fourth, good pooled calibration hid two-fold under-prediction at the hospital with a restricted laboratory menu, which a local intercept removed without sharing data.

### 5.2 What the ledger adds, and what it does not

Most of BFE’s protection comes from two mechanisms that need no blockchain, a median-of-peers gate and a digest comparison; a trusted server or a signed append-only log would reproduce every prediction in this study. The ledger adds three properties. The admission rule runs as code that no participant can change after deployment, and each round’s threshold is recorded on-chain; every input to it (artefact digest, signed score, decision) is non-repudiable and replicated at every hospital; and any hospital can recompute the admitted set and the weights from the ledger alone, so a deviating aggregator that re-admits a rejected model or alters a weight is detectable. These properties matter when hospitals do not fully trust the aggregator or each other [68,69].

The guarantees are narrower than they may appear. Scores are self-reported, so a validator can post a plausible false AUROC without running the model; only grossly deviant scores, such as the zeros in A2, are attributable. The contract does not check the global specification against the admission decisions, so a deviating aggregator is detected, not prevented. Recalibration coefficients (and, for BFE-Stack, the meta-learner) are committed by digest but can be reproduced only by refitting on the hospitals’ validation data, so BFE-Stack, with more fitted coefficients, is less auditable than BFE-PW, although it was the more accurate variant. Finally, the attacks that passed the gate (Section 4.4) show that it certifies average discrimination on peers’ validation data, not the absence of harm to a subgroup or to calibration, and it never inspects recalibration. A consortium that relies on BFE should therefore pair the gate with subgroup-level attestation, which the contract could store as further scores, and with robust recalibration.

### 5.3 Comparison with previous work

Ledger-coordinated clinical learning has so far recorded updates, consent and site-level misbehaviour for parametric or linear models [26–31,65]. BFE differs in recording one admission decision per model artefact, computed by the contract from hash-bound peer scores, and in testing substitution and collusion separately from training-time poisoning. Multi-Krum protected FedAvg as well as the gate did against gross flipping, but the gate leaves an auditable record of each decision. One-round ensembling is established [23,24,52], and model-level federation of random forests has been evaluated across real hospitals [25,49,53,82]; our accuracy results agree with that work, including the findings that small hospitals gain most [49,50] and that decentralised models match central ones in AUROC but lose on threshold-based measures [48].

### 5.4 Clinical deployment and generalisability

A deployment would follow the roles in Fig. 1: one validating node per hospital, a governance board holding the registration key (preferably as a k-of-n multi-signature), and rounds scheduled by the board, for example quarterly or after a coding or assay change. Before scoring its own patients, each hospital verifies the admitted artefacts against the ledger and fits its own intercept on local validation data. The ledger then records which model version was in use, on what evidence it was admitted and who attested to it, which supports the change control that clinical model governance requires [68].

Three results bear on transferability. The protocol accepts any model family that uses the agreed feature schema. Its advantage over local modelling appeared where hospitals had a few thousand admissions, as small and resource-limited hospitals do, although with about 7–25 deaths per validation split peer AUROCs near τ were noisy, and no attacks were tested in that regime. Hospital C, without lactate, blood-gas, troponin or liver panels, resembles many district hospitals in low- and middle-income countries; there a local intercept, which needs no data sharing, raised the share of deaths identified from 3% to 36%. And on the care-era partition, BFE-Stack had the highest AUROC among the federated methods and local-only models no longer outperformed federation. What does not transfer automatically is the threshold τ = 0.70, which assumes outcomes on which a useful model clearly exceeds it; for harder outcomes the threshold, and the number of validators needed for a stable median, must be set from pilot data.

### 5.5 Limitations

The hospitals were carved from one US academic centre, and their heterogeneity was imposed by rule; the care-era partition adds unengineered shift but still comes from one institution, with one feature vocabulary. Real inter-institutional differences in coding, assays and record systems were absent, and no external validation was performed, for example on eICU-CRD [35]. The ledger ran on a single-process EVM without consensus or node failure, so latencies are simulator timings and gas is the transferable quantity. The contract is a prototype without an attestation quorum, deadlines, commit– reveal of scores, a multi-signature governor or key revocation; it blocks self-attestation by address rather than by hospital, and finalizeRound cost grew about as *K*2.5 between 16 and 32 hospitals, so one call would exceed a 30-milliongas block at about 65 hospitals. A deviating aggregator was argued to be detectable but not tested.

Privacy rests on data locality, not on a formal guaran-tee [4]: here “privacy-preserving” means only that no patient record leaves its hospital. Every hospital receives every peer’s full tree models, whose split thresholds and leaf values encode training data; leaves held at least 20 training rows (random forest) or a Hessian weight of 5 (XGBoost), which limits but does not prevent membership inference [83], and a recent attack reconstructed much of clients’ training data from federated tree models [84]. The federated recalibration shares exact per-site gradients and Hessians, and threshold selection shares per-site counts of true and false positives. No differential privacy, secure aggregation [85] or membership-inference audit was applied, and because a model trained on personal data may itself be personal data under the GDPR, sharing it needs its own legal basis. Random forests were exchanged as Python pickles, which a malicious hospital could use to run code on its peers; a deployment needs a non-executable format. The attacks did not include backdoor triggers or adaptive attacks optimised against the gate [11,12,86]. Comparisons mix aggregation rule and model family; the MLP-member ablation separates them only in part, and lossless federated XGBoost was represented only by its centralised ceiling [22]. Each hospital’s validation split served for early stopping, attestation, recalibration and threshold selection, which may make recalibration slightly optimistic. Finally, the outcome was in-hospital death at a 24-hour landmark only, and several analyses were added after the first results were known (see Reporting).

## 6. Conclusion

A smart contract that admits locally trained tree models by hash-bound peer validation scores gave a consortium a verifiable record of which models entered its mortality model and why, excluded grossly poisoned and substituted models and failed closed under collusion, at a cost of about 0.01 AU-ROC against local models with large hospitals and none when hospitals were small. It did not detect attacks that preserved pooled discrimination, and its recalibration needed a robust, median-based form. Its value is auditable governance rather than accuracy, and it now needs testing on a multi-node permissioned network across independent hospitals.

## Supporting information

BFE_BHTY supplement

AI Use summary

Tripod

## Data Availability

MIMIC-IV v3.1 is available from PhysioNet (https://doi.org/10.13026/kpb9-mt58) to credentialed users who complete the required training and sign the PhysioNet Credentialed Health Data Use Agreement. Under that agreement the authors cannot share patient-level data, derived patient-level data or model predictions. All aggregate results are reported in the article and its supplementary material. The complete analysis code, including the Solidity smart contract, is available at https://github.com/mostakphoenixsoftbd/Blockchain-Enabled-Federated-Ensemble-Framework and archived at Zenodo (https://doi.org/10.5281/zenodo.23947714).

https://github.com/mostakphoenixsoftbd/Blockchain-Enabled-Federated-Ensemble-Framework

## Acknowledgments

We thank the MIT Laboratory for Computational Physiology and Beth Israel Deaconess Medical Center for making the MIMIC-IV database available.

## Use of AI tools

An AI coding assistant was used from September to November 2026 as described in Section 3.9 and Supplementary File S2; it received no patient-level data. No generative-AI images were used; all figures were produced by the authors’ code.

## Author contributions

Mostak Ahmed: conceptualisation, methodology, software, formal analysis, data curation, visualisation, writing – original draft, project administration. Sanjida Islam: validation (clinical interpretation), writing – review and editing. Sabera Tabassum Wahid, Golam Sarwar Khondoker, Golam Rabbi, Rony Md Ariful Islam, Md Mostafizur Rahman, Al Rafy and Md. Masuqul Haque: writing – review and editing. All authors meet the ICMJE authorship criteria, approved the final manuscript and agree to be accountable for all aspects of the work.

## Data availability statement

The data that support the findings of this study are available from PhysioNet: MIMIC-IV version 3.1 (https://doi.org/10.13026/kpb9-mt58), to credentialed users who complete the required training and sign the PhysioNet Credentialed Health Data Use Agreement. Under that agreement the authors cannot share patient-level data, derived patient-level data or model predictions. All aggregate results are reported in the article and its supplementary material.

## Code availability

The complete code is available at https://github.com/mostakphoenixsoftbd/Blockchain-Enabled-Federated-Ensemble-Framework and archived at Zenodo (https://doi.org/10.5281/zenodo.23947714). It covers the whole pipeline, including the Solidity contract, and contains no patient data.

## Ethics

This secondary analysis used the de-identified MIMIC-IV database, whose collection and sharing were approved by the institutional review boards of Beth Israel Deaconess Medical Center and the Massachusetts Institute of Technology with a waiver of informed consent. Access was obtained under the PhysioNet Credentialed Health Data Use Agreement. No further ethical approval was required. Patients and the public were not involved in the design, conduct or reporting of this research.

### Reporting

The study is reported according to TRIPOD+AI, with PROBAST used to consider risk of bias (Supplementary File S3). The analysis plan was not registered and no time-stamped protocol exists, so comparisons between BFE variants are exploratory; the data-scarce regime, care-era partition, attacks A5–A7, recalibration variants and added comparators were defined after the first results were known.

## Supporting information

**S1 File. Supplementary material**. Supplementary Methods, Tables and Figures.

**S2 File. Use of AI tools**.

**S3 File. TRIPOD+AI checklist and PROBAST assessment**.

