## Supplementary material for "A Blockchain-Enabled Federated Ensemble Framework for Privacy-Preserving Mortality Prediction Across Heterogeneous Healthcare Systems": BFE_BHTY supplement

### Supplementary Methods

*S-M1. Extended predictor sets (sensitivity analysis)*

The primary predictors omit bedside and treatment data. After the main analysis, we built an extended set from the MIMIC-IV *icu* module and the *hosp* prescription and microbiology tables, using the same window (6 h before to 24 h after admission) and adding 27 columns (175 in total). From the ICU module, these were:

- ICU time in the window and the type of the first care unit;
- the number of distinct vasoactive agents (noradrenaline, adrenaline, vasopressin, phenylephrine, dopamine, dobutamine) and any propofol or midazolam infusion;
- crystalloid volume, prorated to the part of each infusion inside the window, and charted weight;
- invasive or non-invasive ventilation and dialysis;
- urine output and hourly urine rate.

From the prescription and microbiology tables, these were:

- the number of distinct drugs and of antimicrobials ordered;
- indicators for intravenous antimicrobials, vasopressors, insulin, loop diuretics, corticosteroids and intravenous heparin;
- the number of microbiology specimens and indicators for blood, urine and respiratory cultures.

Only events that started in the window were used. Culture *orders* were used, never their results, and comfort-care medications were excluded. ICU-derived values were set to zero for admissions without ICU time in the window, except urine output and weight, which were left missing. The extended columns were joined onto the same admissions, so the hospital partition, the patient-level splits and the test patients were identical to the primary analysis and every comparison is paired. The Hospital C missingness simulation was applied to laboratory panels only, as in the primary analysis.

A second set added bedside vital signs from ICU charting (`chartevents`) in the same window: the minimum, maximum and mean of heart rate, systolic and mean arterial pressure, respiratory rate, peripheral oxygen saturation and temperature, and the lowest eye, verbal and motor Glasgow Coma Scale components (21 columns, 196 in total). Values outside physiological limits were discarded and Fahrenheit temperatures converted to Celsius. MIMIC-IV charts these values only during ICU stays, so they were missing for admissions without ICU care in the window. All clean-condition methods were retrained with each extended set at $p = 0.85$ over the same five seeds; AUROCs were compared by DeLong's test with Holm adjustment (40 tests per set), and per-site O/E and AUROC were reported separately for admissions with and without ICU care in the window.

*S-M2. Smart-contract functions*

| Function | Caller | Effect | Reverts when |
|---|---|---|---|
| `registerNode(address, siteId)` | governor | admits a hospital node | node already registered |
| `openRound(r, τ)` | aggregator | opens round $r$ with admission threshold $\tau$ (AUROC × $10^4$) | round already exists |
| `submitModel(r, h_m, h_meta, n_train)` | registered node | commits the model digest, the digest of a training manifest (in this prototype the model family and hospital; a deployment would add the feature schema and hyper-parameters) and the training-set size | caller unregistered; round closed; second submission in the round |
| `attest(r, owner, h_eval, s, h_record)` | registered node | records the AUROC $s$ (× $10^4$) that the caller measured for *owner*'s model on its own validation set | round closed; self-attestation; no submission by *owner*; $h_{\text{eval}}$ differs from the committed digest; duplicate attestation; score out of range |

| Function | Caller | Effect | Reverts when |
|---|---|---|---|
| `finalizeRound(r)` | aggregator | computes the median of each model's attestations on-chain and admits the model if it has at least one attestation and the median is $\geq \tau$; emits the decision as an event | round not open |
| `commitGlobal(r, h_global, h_weights)` | aggregator | records the digest of the global specification (admitted members by hospital and model family, weights, calibration or stacking coefficients) once per finalised round; the prototype passes the same digest as h_global and h_weights | round not finalised; already committed |
| `verifyModel(r, node, h)` (view) | anyone | true only if $h$ matches an admitted commitment | — |

The contract cannot verify that a validator actually evaluated the artefact or that the reported AUROC is correct: scores are signed, non-repudiable self-reports, and the median rule is what limits the influence of false ones. `commitGlobal` does not check the specification against the admission decisions; consistency is verified off-chain by recomputing the specification from the ledger. The contract has no attestation quorum beyond one, no deadline, no node removal or key rotation, and it blocks self-attestation by address, not by hospital. Each aggregation uses one round per model family, because a node can submit once per round. In all experiments every model received all $K - 1 = 3$ attestations before finalisation.

*S-M3. Software and hardware*

All code was written in Python 3.14 (XGBoost 3.4.1, scikit-learn 1.9.1, PyTorch 2.12.1, statsmodels 0.15.0) and run on an 8-core CPU workstation. The ledger was a local Ethereum Virtual Machine (py-evm via eth-tester, web3.py 8.0) running a Solidity 0.8.24 contract compiled with the optimiser.

*S-M4. Model, comparator and evaluation details*

**Predictors.** Demographics and admission context: age, sex, race group, language, marital status, insurance, admission type and location, arrival through the emergency department, hour and weekend of admission. Utilisation history: prior hospitalisations and days since last discharge. Comorbidities: twelve Charlson-type indicators from ICD-9/ICD-10 codes of previous admissions. Laboratory: minimum, maximum and mean of 29 tests from 6 h before to 24 h after admission, and the total number of laboratory results. Tree models handled missing values natively.

**Simulated hospitals.** Hospital A (geriatric referral) receives patients aged ≥70 years at first eligible admission. Hospital B's kidney-disease rule uses the 24 h maximum creatinine in any admission or a chronic kidney disease diagnosis; it is a fixed property of the patient that determines only the site and never enters the predictors. Hospital C's restricted menu removes lactate, blood-gas pH, troponin T, albumin, liver enzymes, bilirubin, differential counts and coagulation panels in the first 24 h, and 30% of its remaining laboratory results are deleted completely at random. In the care-era partition, the two most recent MIMIC-IV anchor-year groups were merged so that sites have similar sizes.

**Ensemble members.** XGBoost: histogram-based, at most 600 trees, depth 6, learning rate 0.05, row and column subsampling 0.8, minimum child weight 5, early stopping (patience 50) on validation AUPRC. Random forest: 200 trees, maximum depth 14, minimum leaf size 20, $\sqrt{d}$ features per split. Hyper-parameters were fixed before the experiments and shared by all hospitals.

**Federated recalibration and stacking.** At each Newton–Raphson iteration every hospital returns only the gradient (2 values) and Hessian (2 × 2) of its local log-likelihood for the Platt model; the stacking meta-learner is L2-regularised ($\lambda = 10^{-3}$). The ensemble needs one exchange of models plus a few low-dimensional Newton iterations.

**MLP comparators.** Two hidden layers (128 and 64 units, ReLU, dropout 0.1); 40 communication rounds of one local epoch (Adam, learning rate $10^{-3}$, batch 256); aggregation weighted by training-set size; the round with the lowest federated validation loss was retained. Inputs were log-transformed (laboratory values and counts), standardised with federated statistics (hospitals shared per-column sums, sums of squares and non-missing counts), clipped at ±5 SD and mean-imputed with missingness indicators. Centralised and local MLPs used the same architecture with early stopping (patience 5 epochs, at most 40). FedTrim discards the largest and smallest client value per coordinate; Multi-Krum (f = 1) averages the three clients whose updates are closest to each other. Federated logistic regression is L2-regularised ($\lambda = 10^{-3}$) on the standardised features.

**Tamper test.** A small XGBoost model trained on synthetic data was altered by 200 single-bit flips, 200 one-byte appends and 20 retrainings; this exercises the on-chain digest comparison, whose outcome does not depend on the model.

**Evaluation.** Each configuration was repeated over five seeds (three for $p \in \{0.5, 1.0\}$). The pooled test set was the primary evaluation population, and hospital-specific test sets were used for per-site analyses. Age bands were 18–49, 50–69, 70–84 and ≥85 years. Each hospital's validation split served for early stopping, peer attestation, recalibration and threshold selection; the test split was never used before evaluation.

**Supplementary Table S1.** Predictors (148 columns after one-hot encoding with a fixed vocabulary).

age, came_via_ed, admit_hour, weekend_admit, n_prior_adm, days_since_last_disch, hx_chf, hx_mi, hx_pvd, hx_cvd, hx_dementia, hx_copd, hx_diabetes, hx_ckd, hx_liver, hx_cancer, hx_metastatic, hx_hiv, n_lab_results, creatinine_min, creatinine_max, creatinine_mean, bun_min, bun_max, bun_mean, sodium_min, sodium_max, sodium_mean, potassium_min, potassium_max, potassium_mean, chloride_min, chloride_max, chloride_mean, bicarbonate_min, bicarbonate_max, bicarbonate_mean, anion_gap_min, anion_gap_max, anion_gap_mean, glucose_min, glucose_max, glucose_mean, calcium_min, calcium_max, calcium_mean, magnesium_min, magnesium_max, magnesium_mean, phosphate_min, phosphate_max, phosphate_mean, hemoglobin_min, hemoglobin_max, hemoglobin_mean, hematocrit_min, hematocrit_max, hematocrit_mean, wbc_min, wbc_max, wbc_mean, platelets_min, platelets_max, platelets_mean, rdw_min, rdw_max, rdw_mean, lactate_min, lactate_max, lactate_mean, ph_min, ph_max, ph_mean, bilirubin_min, bilirubin_max, bilirubin_mean, alt_min, alt_max, alt_mean, ast_min, ast_max, ast_mean, alp_min, alp_max, alp_mean, albumin_min, albumin_max, albumin_mean, inr_min, inr_max, inr_mean, pt_min, pt_max, pt_mean, ptt_min, ptt_max, ptt_mean, lymphocytes_pct_min, lymphocytes_pct_max, lymphocytes_pct_mean, neutrophils_pct_min, neutrophils_pct_max, neutrophils_pct_mean, troponin_t_min, troponin_t_max, troponin_t_mean, gender_F, gender_M, admission_type_AMBULATORY OBSERVATION, admission_type_DIRECT EMER., admission_type_DIRECT OBSERVATION, admission_type_ELECTIVE, admission_type_EU OBSERVATION, admission_type_EW EMER., admission_type_OBSERVATION ADMIT, admission_type_SURGICAL SAME DAY ADMISSION, admission_type_URGENT, admission_location_AMBULATORY SURGERY TRANSFER, admission_location_CLINIC REFERRAL, admission_location_EMERGENCY ROOM, admission_location_INFORMATION NOT AVAILABLE, admission_location_INTERNAL TRANSFER TO OR FROM PSYCH, admission_location_PACU, admission_location_PHYSICIAN REFERRAL, admission_location_PROCEDURE SITE, admission_location_TRANSFER FROM HOSPITAL, admission_location_TRANSFER FROM SKILLED NURSING FACILITY, admission_location_UNKNOWN, admission_location_WALK-IN/SELF REFERRAL, insurance_Medicaid, insurance_Medicare, insurance_No charge, insurance_Other, insurance_Private, insurance_UNKNOWN, marital_status_DIVORCED, marital_status_MARRIED, marital_status_SINGLE, marital_status_UNKNOWN, marital_status_WIDOWED, race_group_ASIAN, race_group_BLACK, race_group_HISPANIC, race_group_OTHER, race_group_UNKNOWN, race_group_WHITE, language_english_False, language_english_True.

**Supplementary Table S2.** Heterogeneity of hospital populations by purity (seed 0).

| Purity (p) | Mortality across hospitals, % | Mean age across hospitals, years | Hospital share of admissions, % |
|---|---|---|---|
| 0.0 | 2.32–2.38 | 60.4–60.8 | 24.7–25.2 |
| 0.5 | 1.72–3.32 | 54.0–72.2 | 20.0–28.6 |
| 0.85 | 1.36–3.83 | 50.5–78.5 | 16.8–30.9 |
| 1.0 | 1.23–4.01 | 49.1–81.0 | 15.3–32.0 |

**Supplementary Table S3.** Secondary analysis: per-seed DeLong comparisons of BFE-PW with six methods at $p = 0.85$, Holm-adjusted over the 30 seed-by-comparison tests.

| Comparison | ΔAUROC (mean) | p (largest of 5 seeds) | Seeds significant after Holm (of 5) |
|---|---|---|---|
| BFE-PW vs BFE-Stack | −0.0054 | <0.001 | 5 |
| BFE-PW vs Central-XGB | −0.0128 | <0.001 | 5 |
| BFE-PW vs Ens-Uniform | +0.0016 | 0.011 | 4 |
| BFE-PW vs FedAvg-MLP | +0.0001 | 0.950 | 0 |
| BFE-PW vs FedProx-MLP | −0.0047 | 0.200 | 3 |
| BFE-PW vs Local-XGB | −0.0083 | <0.001 | 5 |

**Supplementary Table S4.** Calibration intercept, slope, integrated calibration index (ICI) and observed-to-expected (O/E) ratio by hospital and pooled (mean over five seeds; ideal 0, 1, 0 and 1).

| Analysis | Method | Hospital | Intercept | Slope | ICI | O/E |
|---|---|---|---|---|---|---|
| Recalibration runs | A7-BFE-PW | A | −2.38 | −3.92 | 0.2669 | 0.13 |
| Recalibration runs | A7-BFE-PW | B | −2.82 | −4.43 | 0.2845 | 0.09 |
| Recalibration runs | A7-BFE-PW | C | −3.73 | −5.85 | 0.3531 | 0.04 |
| Recalibration runs | A7-BFE-PW | D | −3.74 | −4.48 | 0.3465 | 0.04 |
| Recalibration runs | A7-BFE-PW | Pooled | −3.07 | −3.98 | 0.3126 | 0.07 |
| Recalibration runs | A7-BFE-PW-Robust | A | 0.34 | 0.93 | 0.0088 | 1.31 |
| Recalibration runs | A7-BFE-PW-Robust | B | −0.34 | 1.05 | 0.0073 | 0.78 |
| Recalibration runs | A7-BFE-PW-Robust | C | 1.21 | 1.39 | 0.0101 | 3.11 |
| Recalibration runs | A7-BFE-PW-Robust | D | 0.24 | 1.07 | 0.0031 | 1.22 |
| Recalibration runs | A7-BFE-PW-Robust | Pooled | 0.26 | 0.94 | 0.0047 | 1.23 |
| Recalibration runs | BFE-PW | A | 0.06 | 0.99 | 0.0035 | 1.05 |
| Recalibration runs | BFE-PW | B | −0.58 | 1.12 | 0.0141 | 0.64 |
| Recalibration runs | BFE-PW | C | 0.8 | 1.48 | 0.0087 | 2.13 |
| Recalibration runs | BFE-PW | D | −0.07 | 1.14 | 0.0022 | 0.95 |
| Recalibration runs | BFE-PW | Pooled | −0.03 | 1.01 | 0.0024 | 0.98 |
| Recalibration runs | BFE-PW-Robust | A | 0.24 | 0.87 | 0.0078 | 1.2 |
| Recalibration runs | BFE-PW-Robust | B | −0.47 | 0.99 | 0.0103 | 0.71 |
| Recalibration runs | BFE-PW-Robust | C | 1.23 | 1.31 | 0.01 | 3.12 |
| Recalibration runs | BFE-PW-Robust | D | 0.17 | 1.01 | 0.0023 | 1.15 |
| Recalibration runs | BFE-PW-Robust | Pooled | 0.17 | 0.89 | 0.0047 | 1.14 |
| Recalibration runs | BFE-PW-SiteInt | A | −0.02 | 0.99 | 0.0032 | 0.99 |
| Recalibration runs | BFE-PW-SiteInt | B | −0.03 | 1.12 | 0.0035 | 0.98 |

| Analysis | Method | Hospital | Intercept | Slope | ICI | O/E |
|---|---|---|---|---|---|---|
| Recalibration runs | BFE-PW-SiteInt | C | −0.08 | 1.48 | 0.0061 | 0.93 |
| Recalibration runs | BFE-PW-SiteInt | D | −0.01 | 1.14 | 0.0025 | 1.0 |
| Recalibration runs | BFE-PW-SiteInt | Pooled | −0.03 | 1.09 | 0.0024 | 0.98 |
| Recalibration runs | FedLR | A | −0.02 | 0.98 | 0.0044 | 0.99 |
| Recalibration runs | FedLR | B | −0.19 | 1.03 | 0.0046 | 0.87 |
| Recalibration runs | FedLR | C | 0.18 | 1.37 | 0.0053 | 1.19 |
| Recalibration runs | FedLR | D | −0.02 | 1.1 | 0.0036 | 0.98 |
| Recalibration runs | FedLR | Pooled | −0.02 | 1.03 | 0.0036 | 0.98 |
| Main runs | BFE-PW | A | 0.06 | 0.99 | 0.0035 | 1.05 |
| Main runs | BFE-PW | B | −0.58 | 1.12 | 0.0141 | 0.64 |
| Main runs | BFE-PW | C | 0.8 | 1.48 | 0.0087 | 2.13 |
| Main runs | BFE-PW | D | −0.07 | 1.14 | 0.0022 | 0.95 |
| Main runs | BFE-PW | Pooled | −0.03 | 1.01 | 0.0024 | 0.98 |
| Main runs | BFE-Stack | A | −0.02 | 0.99 | 0.0036 | 0.98 |
| Main runs | BFE-Stack | B | −0.44 | 1.08 | 0.0098 | 0.72 |
| Main runs | BFE-Stack | C | 0.65 | 1.34 | 0.0074 | 1.82 |
| Main runs | BFE-Stack | D | −0.0 | 1.11 | 0.0023 | 1.0 |
| Main runs | BFE-Stack | Pooled | −0.03 | 1.01 | 0.0023 | 0.98 |
| Main runs | Central-XGB | A | 0.04 | 0.96 | 0.0034 | 1.03 |
| Main runs | Central-XGB | B | −0.17 | 1.01 | 0.0039 | 0.89 |
| Main runs | Central-XGB | C | 0.09 | 1.05 | 0.0028 | 1.09 |
| Main runs | Central-XGB | D | 0.1 | 1.03 | 0.0021 | 1.08 |
| Main runs | Central-XGB | Pooled | 0.02 | 0.99 | 0.0022 | 1.01 |
| Main runs | FedAvg-MLP | A | −0.13 | 0.9 | 0.0056 | 0.91 |
| Main runs | FedAvg-MLP | B | −0.43 | 0.95 | 0.0095 | 0.74 |
| Main runs | FedAvg-MLP | C | 0.59 | 0.97 | 0.0058 | 1.71 |
| Main runs | FedAvg-MLP | D | −0.08 | 0.91 | 0.002 | 0.94 |
| Main runs | FedAvg-MLP | Pooled | −0.09 | 0.88 | 0.0033 | 0.93 |
| Main runs | FedMedian-MLP | A | −0.15 | 0.92 | 0.0059 | 0.89 |
| Main runs | FedMedian-MLP | B | −0.45 | 0.97 | 0.0098 | 0.73 |
| Main runs | FedMedian-MLP | C | 0.59 | 0.97 | 0.0058 | 1.7 |
| Main runs | FedMedian-MLP | D | −0.09 | 0.93 | 0.0018 | 0.93 |
| Main runs | FedMedian-MLP | Pooled | −0.11 | 0.9 | 0.0033 | 0.92 |
| Main runs | FedProx-MLP | A | −0.12 | 0.95 | 0.0045 | 0.91 |
| Main runs | FedProx-MLP | B | −0.43 | 0.99 | 0.0092 | 0.74 |
| Main runs | FedProx-MLP | C | 0.5 | 1.04 | 0.0051 | 1.57 |
| Main runs | FedProx-MLP | D | −0.06 | 0.94 | 0.0016 | 0.95 |
| Main runs | FedProx-MLP | Pooled | −0.1 | 0.93 | 0.0026 | 0.93 |
| Main runs | Local-XGB | A | 0.03 | 1.01 | 0.0037 | 1.02 |
| Main runs | Local-XGB | B | −0.01 | 0.99 | 0.0039 | 0.99 |
| Main runs | Local-XGB | C | 0.08 | 1.04 | 0.0026 | 1.07 |
| Main runs | Local-XGB | D | 0.03 | 1.01 | 0.0022 | 1.02 |
| Main runs | Local-XGB | Pooled | 0.03 | 1.01 | 0.0026 | 1.02 |

**Supplementary Table S5.** Joint patient-clustered bootstrap differences for the data-scarce regimes, the care-era partition and the recalibration runs (95% CIs from 1,000 replicates).

| Analysis | Comparison | Metric | Δ (95% CI) | Seeds Δ > 0 |
|---|---|---|---|---|
| Recalibration runs | BFE-PW − FedLR | AUROC | +0.0068 (+0.0051 to +0.0085) | 5 |
| Recalibration runs | BFE-PW − FedLR | AUPRC | +0.0285 (+0.0237 to +0.0337) | 5 |
| Recalibration runs | BFE-Stack − FedLR | AUROC | +0.0122 (+0.0107 to +0.0137) | 5 |
| Recalibration runs | BFE-Stack − FedLR | AUPRC | +0.0369 (+0.0321 to +0.0422) | 5 |
| Care-era sites | BFE-PW − FedAvg-MLP | AUROC | +0.0004 (−0.0009 to +0.0019) | 2 |
| Care-era sites | BFE-PW − FedAvg-MLP | AUPRC | −0.0100 (−0.0145 to −0.0050) | 0 |
| Care-era sites | BFE-PW − FedProx-MLP | AUROC | −0.0018 (−0.0029 to −0.0004) | 1 |
| Care-era sites | BFE-PW − FedProx-MLP | AUPRC | −0.0146 (−0.0189 to −0.0102) | 0 |
| Care-era sites | BFE-PW − Local-XGB | AUROC | +0.0011 (−0.0000 to +0.0021) | 4 |
| Care-era sites | BFE-PW − Local-XGB | AUPRC | +0.0037 (−0.0004 to +0.0080) | 5 |
| Care-era sites | BFE-PW − Central-XGB | AUROC | −0.0059 (−0.0069 to −0.0050) | 0 |
| Care-era sites | BFE-PW − Central-XGB | AUPRC | −0.0248 (−0.0290 to −0.0205) | 0 |
| Care-era sites | BFE-Stack − FedAvg-MLP | AUROC | +0.0043 (+0.0032 to +0.0055) | 5 |
| Care-era sites | BFE-Stack − FedAvg-MLP | AUPRC | +0.0017 (−0.0025 to +0.0062) | 3 |
| Care-era sites | BFE-Stack − FedProx-MLP | AUROC | +0.0021 (+0.0012 to +0.0033) | 5 |
| Care-era sites | BFE-Stack − FedProx-MLP | AUPRC | −0.0030 (−0.0067 to +0.0012) | 2 |
| Care-era sites | BFE-Stack − Local-XGB | AUROC | +0.0050 (+0.0041 to +0.0059) | 5 |
| Care-era sites | BFE-Stack − Local-XGB | AUPRC | +0.0154 (+0.0116 to +0.0193) | 5 |
| Care-era sites | BFE-Stack − Central-XGB | AUROC | −0.0020 (−0.0026 to −0.0013) | 0 |

| Analysis | Comparison | Metric | Δ (95% CI) | Seeds Δ > 0 |
|---|---|---|---|---|
| Care-era sites | BFE-Stack − Central-XGB | AUPRC | −0.0132 (−0.0170 to −0.0096) | 0 |
| 5% of data | BFE-PW − FedAvg-MLP | AUROC | +0.0030 (+0.0005 to +0.0060) | 4 |
| 5% of data | BFE-PW − FedAvg-MLP | AUPRC | −0.0088 (−0.0127 to −0.0042) | 0 |
| 5% of data | BFE-PW − FedProx-MLP | AUROC | −0.0014 (−0.0040 to +0.0015) | 1 |
| 5% of data | BFE-PW − FedProx-MLP | AUPRC | −0.0160 (−0.0200 to −0.0114) | 0 |
| 5% of data | BFE-PW − Local-XGB | AUROC | +0.0219 (+0.0189 to +0.0249) | 5 |
| 5% of data | BFE-PW − Local-XGB | AUPRC | +0.0280 (+0.0223 to +0.0331) | 5 |
| 5% of data | BFE-PW − Central-XGB | AUROC | −0.0172 (−0.0192 to −0.0151) | 1 |
| 5% of data | BFE-PW − Central-XGB | AUPRC | −0.0276 (−0.0324 to −0.0228) | 0 |
| 5% of data | BFE-Stack − FedAvg-MLP | AUROC | +0.0029 (+0.0004 to +0.0058) | 4 |
| 5% of data | BFE-Stack − FedAvg-MLP | AUPRC | −0.0133 (−0.0176 to −0.0082) | 1 |
| 5% of data | BFE-Stack − FedProx-MLP | AUROC | −0.0015 (−0.0042 to +0.0014) | 1 |
| 5% of data | BFE-Stack − FedProx-MLP | AUPRC | −0.0204 (−0.0248 to −0.0154) | 0 |
| 5% of data | BFE-Stack − Local-XGB | AUROC | +0.0218 (+0.0188 to +0.0248) | 5 |
| 5% of data | BFE-Stack − Local-XGB | AUPRC | +0.0235 (+0.0185 to +0.0283) | 4 |
| 5% of data | BFE-Stack − Central-XGB | AUROC | −0.0173 (−0.0193 to −0.0153) | 0 |
| 5% of data | BFE-Stack − Central-XGB | AUPRC | −0.0321 (−0.0373 to −0.0275) | 0 |
| 10% of data | BFE-PW − FedAvg-MLP | AUROC | +0.0019 (−0.0004 to +0.0045) | 3 |
| 10% of data | BFE-PW − FedAvg-MLP | AUPRC | −0.0122 (−0.0160 to −0.0078) | 0 |
| 10% of data | BFE-PW − FedProx-MLP | AUROC | −0.0021 (−0.0043 to +0.0004) | 1 |
| 10% of data | BFE-PW − FedProx-MLP | AUPRC | −0.0214 (−0.0255 to −0.0170) | 0 |
| 10% of data | BFE-PW − Local-XGB | AUROC | +0.0022 (+0.0001 to +0.0044) | 4 |
| 10% of data | BFE-PW − Local-XGB | AUPRC | +0.0020 (−0.0027 to +0.0066) | 3 |
| 10% of data | BFE-PW − Central-XGB | AUROC | −0.0188 (−0.0207 to −0.0169) | 0 |
| 10% of data | BFE-PW − Central-XGB | AUPRC | −0.0395 (−0.0445 to −0.0343) | 0 |
| 10% of data | BFE-Stack − FedAvg-MLP | AUROC | +0.0037 (+0.0015 to +0.0063) | 5 |
| 10% of data | BFE-Stack − FedAvg-MLP | AUPRC | −0.0073 (−0.0117 to −0.0023) | 1 |
| 10% of data | BFE-Stack − FedProx-MLP | AUROC | −0.0003 (−0.0025 to +0.0021) | 2 |
| 10% of data | BFE-Stack − FedProx-MLP | AUPRC | −0.0165 (−0.0210 to −0.0115) | 0 |
| 10% of data | BFE-Stack − Local-XGB | AUROC | +0.0040 (+0.0018 to +0.0062) | 3 |
| 10% of data | BFE-Stack − Local-XGB | AUPRC | +0.0069 (+0.0023 to +0.0111) | 5 |
| 10% of data | BFE-Stack − Central-XGB | AUROC | −0.0170 (−0.0187 to −0.0152) | 0 |
| 10% of data | BFE-Stack − Central-XGB | AUPRC | −0.0346 (−0.0395 to −0.0293) | 0 |
| 20% of data | BFE-PW − FedAvg-MLP | AUROC | +0.0009 (−0.0011 to +0.0031) | 4 |
| 20% of data | BFE-PW − FedAvg-MLP | AUPRC | −0.0097 (−0.0140 to −0.0052) | 0 |
| 20% of data | BFE-PW − FedProx-MLP | AUROC | −0.0045 (−0.0064 to −0.0025) | 0 |
| 20% of data | BFE-PW − FedProx-MLP | AUPRC | −0.0272 (−0.0321 to −0.0221) | 0 |
| 20% of data | BFE-PW − Local-XGB | AUROC | −0.0025 (−0.0044 to −0.0007) | 2 |
| 20% of data | BFE-PW − Local-XGB | AUPRC | −0.0042 (−0.0091 to +0.0001) | 1 |
| 20% of data | BFE-PW − Central-XGB | AUROC | −0.0170 (−0.0184 to −0.0154) | 0 |
| 20% of data | BFE-PW − Central-XGB | AUPRC | −0.0396 (−0.0442 to −0.0348) | 0 |
| 20% of data | BFE-Stack − FedAvg-MLP | AUROC | +0.0030 (+0.0011 to +0.0050) | 5 |
| 20% of data | BFE-Stack − FedAvg-MLP | AUPRC | −0.0051 (−0.0094 to −0.0003) | 1 |
| 20% of data | BFE-Stack − FedProx-MLP | AUROC | −0.0024 (−0.0042 to −0.0005) | 1 |
| 20% of data | BFE-Stack − FedProx-MLP | AUPRC | −0.0226 (−0.0276 to −0.0172) | 0 |
| 20% of data | BFE-Stack − Local-XGB | AUROC | −0.0004 (−0.0022 to +0.0014) | 2 |

| Analysis | Comparison | Metric | Δ (95% CI) | Seeds Δ > 0 |
|---|---|---|---|---|
| 20% of data | BFE-Stack − Local-XGB | AUPRC | +0.0005 (−0.0045 to +0.0048) | 2 |
| 20% of data | BFE-Stack − Central-XGB | AUROC | −0.0149 (−0.0162 to −0.0134) | 0 |
| 20% of data | BFE-Stack − Central-XGB | AUPRC | −0.0350 (−0.0394 to −0.0303) | 0 |

**Supplementary Table S6.** Number of the attacker's two members admitted by the contract in each seed, by scenario.

| Scenario | Attacker members admitted (of 2), per seed |
|---|---|
| Clean | 2 2 2 2 2 |
| A1 label flip | 0 0 0 0 0 |
| A2 flip + false attestation | 0 0 0 0 0 |
| A4 collusion (C and D) | 0 0 0 0 0 |
| A5 partial flip 20% | 2 2 2 2 2 |
| A5 partial flip 40% | 0 0 1 0 0 |
| A6 targeted flip (female deaths) | 2 2 2 2 2 |

**Supplementary Table S7.** Targeted attack A6: AUROC, sensitivity and positive predictive value among female and male patients (mean ± SD over five seeds).

| Model | Sex | AUROC | Sensitivity | PPV |
|---|---|---|---|---|
| Clean BFE-PW | Female | 0.915 ± 0.005 | 0.328 ± 0.027 | 0.363 ± 0.031 |
| Clean BFE-PW | Male | 0.887 ± 0.006 | 0.351 ± 0.038 | 0.334 ± 0.029 |
| A6 BFE-PW | Female | 0.913 ± 0.005 | 0.275 ± 0.034 | 0.413 ± 0.047 |
| A6 BFE-PW | Male | 0.886 ± 0.007 | 0.439 ± 0.036 | 0.256 ± 0.032 |
| Oracle BFE-PW | Female | 0.916 ± 0.005 | 0.331 ± 0.031 | 0.358 ± 0.037 |
| Oracle BFE-PW | Male | 0.887 ± 0.006 | 0.354 ± 0.040 | 0.328 ± 0.029 |
| Clean FedAvg | Female | 0.914 ± 0.006 | 0.356 ± 0.053 | 0.358 ± 0.033 |
| Clean FedAvg | Male | 0.888 ± 0.005 | 0.363 ± 0.058 | 0.350 ± 0.054 |
| A6 FedAvg | Female | 0.909 ± 0.005 | 0.292 ± 0.018 | 0.404 ± 0.024 |
| A6 FedAvg | Male | 0.885 ± 0.004 | 0.457 ± 0.026 | 0.260 ± 0.022 |

**Supplementary Table S8.** Subgroup audit on the pooled test set ($p = 0.85$; means over five seeds; admissions and deaths are per-seed means). Strata with fewer than 20 deaths were not analysed. O/E, observed-to-expected deaths.

| Attribute | Group | Method | Admissions | Deaths | AUROC (SD) | AUPRC | O/E |
|---|---|---|---|---|---|---|---|
| Age | 18–49 | BFE-PW | 23,336 | 166 | 0.957 (0.010) | 0.365 | 0.76 |
| Age | 18–49 | BFE-Stack | 23,336 | 166 | 0.959 (0.010) | 0.380 | 0.95 |
| Age | 18–49 | Central-XGB | 23,336 | 166 | 0.965 (0.007) | 0.416 | 1.00 |
| Age | 18–49 | FedAvg-MLP | 23,336 | 166 | 0.951 (0.009) | 0.382 | 1.03 |
| Age | 18–49 | FedProx-MLP | 23,336 | 166 | 0.954 (0.008) | 0.390 | 1.01 |
| Age | 18–49 | Local-XGB | 23,336 | 166 | 0.962 (0.009) | 0.373 | 0.96 |
| Age | 50–69 | BFE-PW | 31,818 | 646 | 0.894 (0.008) | 0.280 | 0.93 |
| Age | 50–69 | BFE-Stack | 31,818 | 646 | 0.902 (0.008) | 0.292 | 0.97 |
| Age | 50–69 | Central-XGB | 31,818 | 646 | 0.913 (0.007) | 0.334 | 1.00 |
| Age | 50–69 | FedAvg-MLP | 31,818 | 646 | 0.895 (0.008) | 0.301 | 0.94 |
| Age | 50–69 | FedProx-MLP | 31,818 | 646 | 0.903 (0.008) | 0.309 | 0.94 |
| Age | 50–69 | Local-XGB | 31,818 | 646 | 0.909 (0.009) | 0.304 | 1.02 |
| Age | 70–84 | BFE-PW | 21,405 | 714 | 0.868 (0.006) | 0.316 | 1.00 |
| Age | 70–84 | BFE-Stack | 21,405 | 714 | 0.872 (0.008) | 0.319 | 0.96 |
| Age | 70–84 | Central-XGB | 21,405 | 714 | 0.878 (0.006) | 0.341 | 1.02 |
| Age | 70–84 | FedAvg-MLP | 21,405 | 714 | 0.867 (0.007) | 0.326 | 0.88 |
| Age | 70–84 | FedProx-MLP | 21,405 | 714 | 0.871 (0.007) | 0.333 | 0.88 |
| Age | 70–84 | Local-XGB | 21,405 | 714 | 0.872 (0.007) | 0.324 | 1.02 |
| Age | ≥85 | BFE-PW | 8,667 | 430 | 0.832 (0.002) | 0.294 | 1.17 |
| Age | ≥85 | BFE-Stack | 8,667 | 430 | 0.837 (0.006) | 0.299 | 1.04 |
| Age | ≥85 | Central-XGB | 8,667 | 430 | 0.841 (0.005) | 0.304 | 1.03 |
| Age | ≥85 | FedAvg-MLP | 8,667 | 430 | 0.824 (0.005) | 0.291 | 0.98 |
| Age | ≥85 | FedProx-MLP | 8,667 | 430 | 0.831 (0.005) | 0.300 | 0.96 |
| Age | ≥85 | Local-XGB | 8,667 | 430 | 0.835 (0.006) | 0.299 | 1.06 |
| Race | Asian | BFE-PW | 2,999 | 71 | 0.905 (0.008) | 0.279 | 1.06 |
| Race | Asian | BFE-Stack | 2,999 | 71 | 0.907 (0.008) | 0.288 | 1.06 |
| Race | Asian | Central-XGB | 2,999 | 71 | 0.914 (0.008) | 0.314 | 1.05 |
| Race | Asian | FedAvg-MLP | 2,999 | 71 | 0.906 (0.011) | 0.286 | 0.97 |
| Race | Asian | FedProx-MLP | 2,999 | 71 | 0.907 (0.005) | 0.296 | 0.94 |
| Race | Asian | Local-XGB | 2,999 | 71 | 0.910 (0.013) | 0.285 | 1.11 |
| Race | Black | BFE-PW | 13,248 | 205 | 0.900 (0.012) | 0.230 | 0.81 |
| Race | Black | BFE-Stack | 13,248 | 205 | 0.907 (0.010) | 0.236 | 0.89 |
| Race | Black | Central-XGB | 13,248 | 205 | 0.916 (0.009) | 0.261 | 1.01 |
| Race | Black | FedAvg-MLP | 13,248 | 205 | 0.900 (0.009) | 0.255 | 0.93 |

| Attribute | Group | Method | Admissions | Deaths | AUROC (SD) | AUPRC | O/E |
|---|---|---|---|---|---|---|---|
| Race | Black | FedProx-MLP | 13,248 | 205 | 0.906 (0.006) | 0.254 | 0.90 |
| Race | Black | Local-XGB | 13,248 | 205 | 0.909 (0.007) | 0.244 | 0.98 |
| Race | Hispanic | BFE-PW | 4,605 | 59 | 0.902 (0.018) | 0.239 | 0.83 |
| Race | Hispanic | BFE-Stack | 4,605 | 59 | 0.907 (0.016) | 0.241 | 0.93 |
| Race | Hispanic | Central-XGB | 4,605 | 59 | 0.914 (0.015) | 0.249 | 1.05 |
| Race | Hispanic | FedAvg-MLP | 4,605 | 59 | 0.903 (0.016) | 0.258 | 0.99 |
| Race | Hispanic | FedProx-MLP | 4,605 | 59 | 0.907 (0.011) | 0.265 | 0.96 |
| Race | Hispanic | Local-XGB | 4,605 | 59 | 0.911 (0.016) | 0.240 | 0.98 |
| Race | Other | BFE-PW | 3,842 | 72 | 0.919 (0.009) | 0.323 | 0.93 |
| Race | Other | BFE-Stack | 3,842 | 72 | 0.923 (0.009) | 0.337 | 0.97 |
| Race | Other | Central-XGB | 3,842 | 72 | 0.927 (0.014) | 0.362 | 1.00 |
| Race | Other | FedAvg-MLP | 3,842 | 72 | 0.918 (0.006) | 0.346 | 0.93 |
| Race | Other | FedProx-MLP | 3,842 | 72 | 0.923 (0.007) | 0.349 | 0.90 |
| Race | Other | Local-XGB | 3,842 | 72 | 0.921 (0.016) | 0.324 | 1.00 |
| Race | Unknown | BFE-PW | 3,448 | 309 | 0.888 (0.005) | 0.498 | 1.28 |
| Race | Unknown | BFE-Stack | 3,448 | 309 | 0.894 (0.005) | 0.505 | 1.15 |
| Race | Unknown | Central-XGB | 3,448 | 309 | 0.906 (0.006) | 0.541 | 1.06 |
| Race | Unknown | FedAvg-MLP | 3,448 | 309 | 0.886 (0.007) | 0.508 | 0.99 |
| Race | Unknown | FedProx-MLP | 3,448 | 309 | 0.895 (0.005) | 0.519 | 1.02 |
| Race | Unknown | Local-XGB | 3,448 | 309 | 0.899 (0.003) | 0.507 | 1.14 |
| Race | White | BFE-PW | 57,084 | 1,239 | 0.892 (0.005) | 0.270 | 0.96 |
| Race | White | BFE-Stack | 57,084 | 1,239 | 0.898 (0.005) | 0.276 | 0.96 |
| Race | White | Central-XGB | 57,084 | 1,239 | 0.905 (0.005) | 0.299 | 1.00 |
| Race | White | FedAvg-MLP | 57,084 | 1,239 | 0.892 (0.004) | 0.278 | 0.92 |
| Race | White | FedProx-MLP | 57,084 | 1,239 | 0.897 (0.004) | 0.286 | 0.91 |
| Race | White | Local-XGB | 57,084 | 1,239 | 0.901 (0.006) | 0.282 | 1.00 |
| Sex | Female | BFE-PW | 44,999 | 902 | 0.915 (0.005) | 0.299 | 1.00 |
| Sex | Female | BFE-Stack | 44,999 | 902 | 0.920 (0.005) | 0.308 | 0.98 |
| Sex | Female | Central-XGB | 44,999 | 902 | 0.926 (0.004) | 0.336 | 1.01 |
| Sex | Female | FedAvg-MLP | 44,999 | 902 | 0.914 (0.006) | 0.311 | 0.95 |
| Sex | Female | FedProx-MLP | 44,999 | 902 | 0.919 (0.006) | 0.321 | 0.94 |
| Sex | Female | Local-XGB | 44,999 | 902 | 0.921 (0.005) | 0.311 | 1.01 |
| Sex | Male | BFE-PW | 40,228 | 1,054 | 0.887 (0.006) | 0.305 | 0.96 |
| Sex | Male | BFE-Stack | 40,228 | 1,054 | 0.893 (0.007) | 0.313 | 0.98 |
| Sex | Male | Central-XGB | 40,228 | 1,054 | 0.902 (0.006) | 0.339 | 1.02 |
| Sex | Male | FedAvg-MLP | 40,228 | 1,054 | 0.888 (0.005) | 0.319 | 0.92 |
| Sex | Male | FedProx-MLP | 40,228 | 1,054 | 0.892 (0.005) | 0.325 | 0.92 |
| Sex | Male | Local-XGB | 40,228 | 1,054 | 0.898 (0.006) | 0.320 | 1.03 |

**Supplementary Table S9.** Ledger operations per experiment (primary configuration; counts include the MLP-member ablation and the attack scenarios). Latency is local-EVM time.

| Operation | Calls per run | Gas (mean) | Latency ms (mean) |
|---|---|---|---|
| deploy | 1 | 1147766 | 191.6 |
| registerNode | 4 | 77538 | 98.2 |
| openRound | 13 | 47959 | 92.8 |
| submitModel | 52 | 127584 | 105.1 |
| attest | 156 | 78655 | 127.5 |
| finalizeRound | 13 | 102853 | 285.4 |
| commitGlobal | 6 | 72905 | 118.1 |

**Supplementary Table S10.** Pooled-test AUROC and AUPRC by skew purity (means and SDs over seeds).

| Purity (p) | Method | AUROC | AUPRC | Seeds |
|---|---|---|---|---|
| 0.0 | Central-XGB | 0.912 ± 0.003 | 0.325 ± 0.006 | 5 |
| 0.0 | BFE-Stack | 0.906 ± 0.002 | 0.297 ± 0.011 | 5 |
| 0.0 | BFE-PW | 0.900 ± 0.002 | 0.286 ± 0.011 | 5 |
| 0.0 | Local-XGB | 0.905 ± 0.004 | 0.300 ± 0.012 | 5 |
| 0.0 | FedAvg-MLP | 0.897 ± 0.003 | 0.294 ± 0.010 | 5 |
| 0.0 | FedProx-MLP | 0.904 ± 0.003 | 0.304 ± 0.009 | 5 |
| 0.0 | Ens-Uniform | 0.901 ± 0.003 | 0.291 ± 0.012 | 5 |
| 0.5 | Central-XGB | 0.915 ± 0.003 | 0.337 ± 0.008 | 3 |
| 0.5 | BFE-Stack | 0.907 ± 0.003 | 0.308 ± 0.006 | 3 |
| 0.5 | BFE-PW | 0.903 ± 0.002 | 0.297 ± 0.007 | 3 |
| 0.5 | Local-XGB | 0.907 ± 0.001 | 0.308 ± 0.007 | 3 |
| 0.5 | FedAvg-MLP | 0.901 ± 0.002 | 0.307 ± 0.002 | 3 |
| 0.5 | FedProx-MLP | 0.907 ± 0.003 | 0.313 ± 0.004 | 3 |
| 0.5 | Ens-Uniform | 0.903 ± 0.003 | 0.299 ± 0.006 | 3 |
| 0.85 | Central-XGB | 0.915 ± 0.003 | 0.337 ± 0.012 | 5 |
| 0.85 | BFE-Stack | 0.907 ± 0.004 | 0.309 ± 0.008 | 5 |
| 0.85 | BFE-PW | 0.902 ± 0.004 | 0.301 ± 0.010 | 5 |
| 0.85 | Local-XGB | 0.910 ± 0.004 | 0.315 ± 0.011 | 5 |
| 0.85 | FedAvg-MLP | 0.902 ± 0.003 | 0.315 ± 0.012 | 5 |

| Purity (p) | Method | AUROC | AUPRC | Seeds |
|---|---|---|---|---|
| 0.85 | FedProx-MLP | 0.906 ± 0.003 | 0.322 ± 0.011 | 5 |
| 0.85 | Ens-Uniform | 0.900 ± 0.004 | 0.303 ± 0.010 | 5 |
| 1.0 | Central-XGB | 0.917 ± 0.002 | 0.343 ± 0.007 | 3 |
| 1.0 | BFE-Stack | 0.907 ± 0.001 | 0.316 ± 0.004 | 3 |
| 1.0 | BFE-PW | 0.896 ± 0.002 | 0.292 ± 0.006 | 3 |
| 1.0 | Local-XGB | 0.913 ± 0.002 | 0.326 ± 0.008 | 3 |
| 1.0 | FedAvg-MLP | 0.903 ± 0.003 | 0.319 ± 0.002 | 3 |
| 1.0 | FedProx-MLP | 0.909 ± 0.002 | 0.327 ± 0.003 | 3 |
| 1.0 | Ens-Uniform | 0.892 ± 0.004 | 0.292 ± 0.007 | 3 |

**Supplementary Table S11.** Sensitivity analysis with extended predictor sets (Supplementary Methods S-M1): pooled-test performance on the same partitions and splits ($p = 0.85$; mean ± SD over five seeds; Δ paired by seed).

| Method | Predictors | AUROC | ΔAUROC vs primary | AUPRC | ΔAUPRC vs primary |
|---|---|---|---|---|---|
| Central-XGB | Extended | 0.925 ± 0.003 | +0.010 | 0.370 ± 0.009 | +0.034 |
| Local-XGB | Extended | 0.919 ± 0.003 | +0.009 | 0.347 ± 0.010 | +0.032 |
| FedAvg-MLP | Extended | 0.914 ± 0.003 | +0.013 | 0.354 ± 0.009 | +0.039 |
| FedProx-MLP | Extended | 0.918 ± 0.004 | +0.012 | 0.360 ± 0.009 | +0.038 |
| Ens-Uniform | Extended | 0.912 ± 0.003 | +0.011 | 0.336 ± 0.010 | +0.033 |
| BFE-PW | Extended | 0.914 ± 0.003 | +0.013 | 0.335 ± 0.010 | +0.035 |
| BFE-Stack | Extended | 0.920 ± 0.003 | +0.012 | 0.351 ± 0.007 | +0.041 |
| Central-XGB | Extended + vital signs | 0.928 ± 0.003 | +0.014 | 0.404 ± 0.004 | +0.068 |
| Local-XGB | Extended + vital signs | 0.923 ± 0.004 | +0.013 | 0.379 ± 0.009 | +0.064 |
| FedAvg-MLP | Extended + vital signs | 0.919 ± 0.003 | +0.018 | 0.386 ± 0.010 | +0.071 |
| FedProx-MLP | Extended + vital signs | 0.922 ± 0.003 | +0.016 | 0.396 ± 0.008 | +0.073 |
| Ens-Uniform | Extended + vital signs | 0.915 ± 0.003 | +0.015 | 0.370 ± 0.009 | +0.067 |
| BFE-PW | Extended + vital signs | 0.918 ± 0.003 | +0.016 | 0.371 ± 0.009 | +0.070 |
| BFE-Stack | Extended + vital signs | 0.924 ± 0.003 | +0.017 | 0.388 ± 0.006 | +0.079 |

**Supplementary Fig. S1.** Participant flow. MIMIC-IV v3.1 admissions: 546,028 (223,452 patients) → excluded, age <18 years: 0 → excluded, length of stay <24 h: 120,812 → excluded, death within 24 h: 1 → analysis cohort: 425,215 admissions, 186,103 patients, 9,984 deaths (2.35%) → patient-level partition into four hospitals and 70/10/20 training/validation/test splits within each hospital.

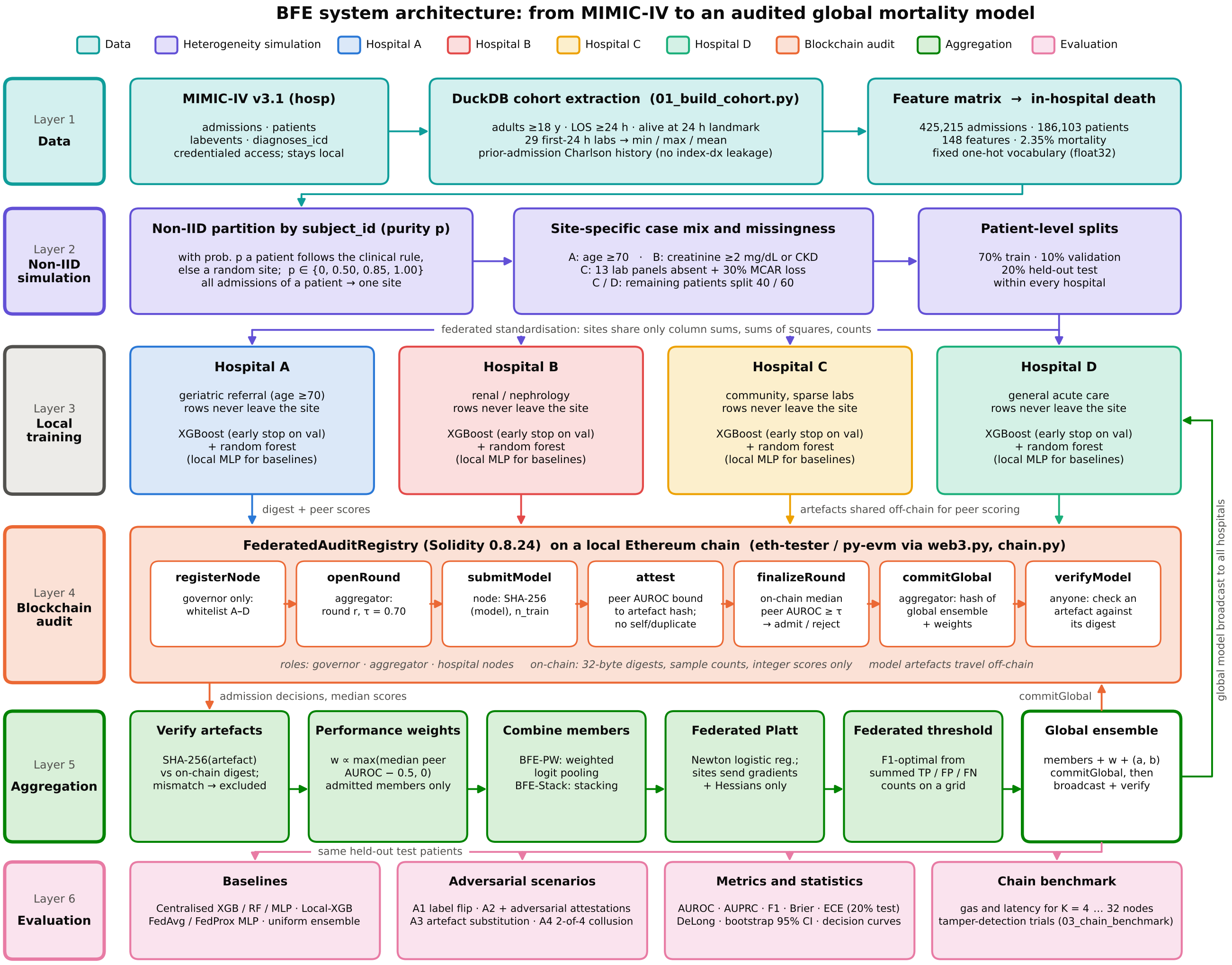


**Supplementary Fig. S2.** End-to-end system architecture.

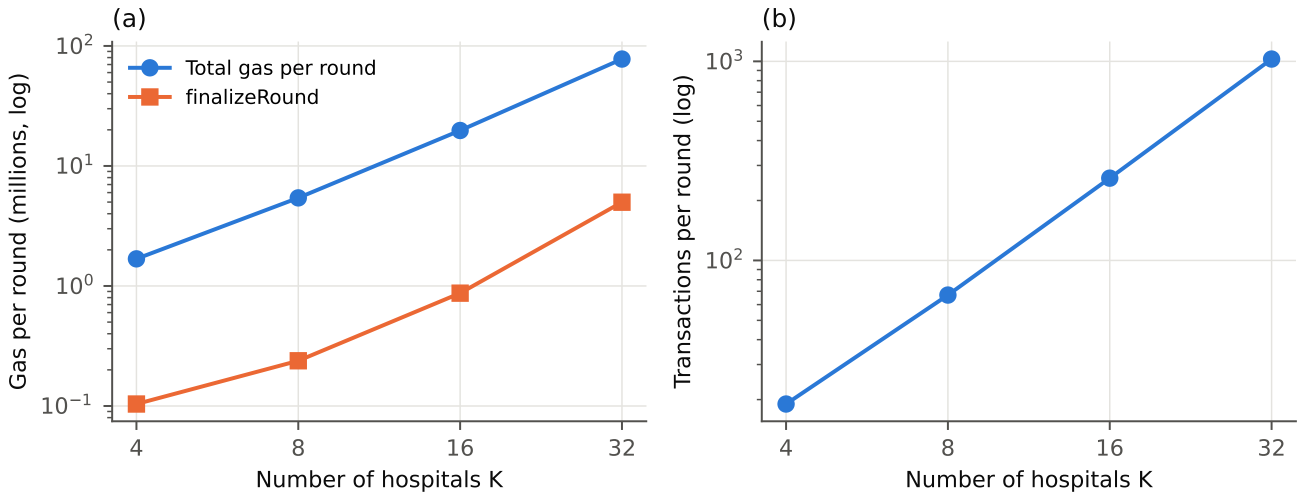


**Supplementary Fig. S3.** Ledger scalability on the local EVM (log–log axes): (a) total gas per round and gas of `finalizeRound`; (b) transactions per round, against the number of hospitals $K$.

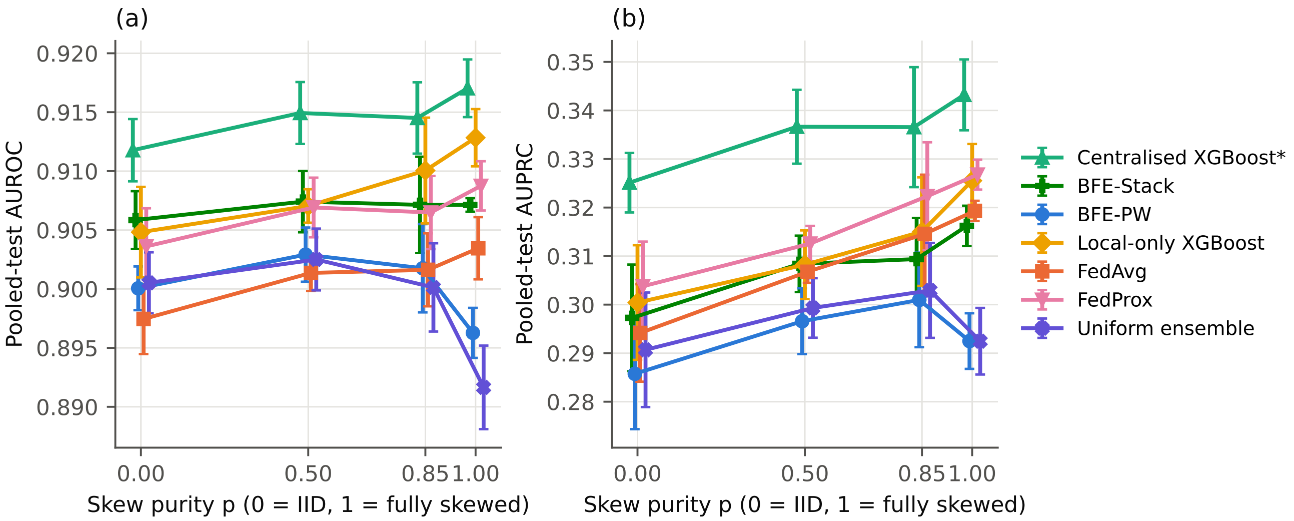


**Supplementary Fig. S4.** Pooled-test (a) AUROC and (b) AUPRC as skew purity increases from IID ($p$ = 0) to fully deterministic referral ($p$ = 1); mean ± SD over five ($p$ = 0, 0.85) or three ($p$ = 0.5, 1.0) seeds.

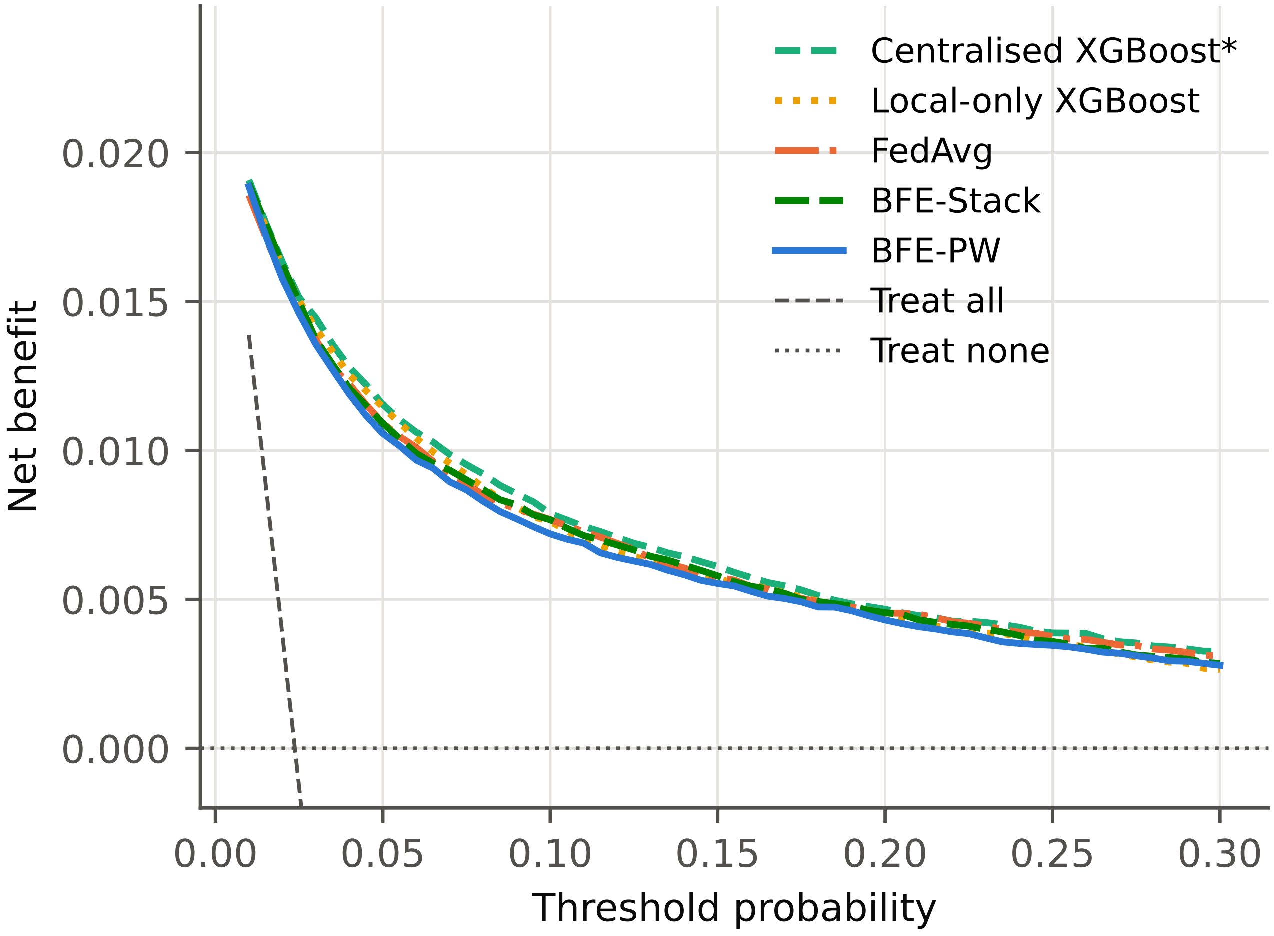


**Supplementary Fig. S5.** Decision-curve analysis of the pooled test set (seed 0, $p$ = 0.85).
