## Supplementary material for "A Blockchain-Enabled Federated Ensemble Framework for Privacy-Preserving Mortality Prediction Across Heterogeneous Healthcare Systems": AI Use summary

### Supplementary File S2. Use of AI assistants in this study

Journal policy (WAME recommendations, as adopted by Blockchain in Healthcare Today) asks authors to describe how AI tools were used and to specify the prompts used to generate text, tables, figures or code. This file summarises that use.

#### Tools

An AI coding assistant with agentic capability (a large-language-model tool), run on the authors’ workstation, where it read and wrote project files and ran commands with the authors’ permission. It was used from September to November 2026. Some sub-tasks (literature checks, reference verification, figure redrawing, manuscript review) were delegated to sub-agents of the same tool under the same instructions. The assistant received code, aggregate results and log summaries only; patient-level MIMIC-IV data never left the workstation, in accordance with the PhysioNet data use agreement.

#### What the assistant did

| Task | Assistant’s role | Authors’ role |
| --- | --- | --- |
| Study design | Proposed the protocol, partitions, comparators and attack scenarios in response to the authors’ brief | Set the research question and target journal; approved the design and every change |
| Data extraction and code | Wrote all Python, SQL and Solidity code (cohort extraction, partitions, models, contract, attacks, statistics, figures) | Provided credentialed data access on their own machine; approved runs |
| Analysis | Ran the analyses locally; no patient-level data left the authors’ computer | Reviewed the outputs |
| Literature | Searched PubMed, Crossref, arXiv and publisher pages; verified every reference’s metadata | Checked and approved the reference list |
| Writing | Drafted and revised all sections, tables, figure captions and supplementary material | Reviewed, edited and approved the text |
| Quality control | Audited every number in the text against the result files; checked rendered pages and figures | Independent review in a separate AI workspace and by the co-authors |

#### Main instructions given to the assistant (paraphrased, in order)

1. Prepare a manuscript for a peer-reviewed journal, “A Blockchain-Enabled Federated Ensemble Framework for Privacy-Preserving Mortality Prediction Across Heterogeneous Healthcare Systems”, for submission as PDF and DOCX, using MIMIC-IV data on the authors’ computer.
2. Use only real experiments and verified citations; report honestly where the proposed method does not outperform its comparators.
3. Build multicolour architecture diagrams, and correct factual errors in the authors’ own mock-ups.
4. Add extended predictors (ICU, medications, microbiology, bedside vital signs) once the full chartevents table was downloaded, and report them as a sensitivity analysis.
5. Review the whole project before submission (literature, methods, gaps, novelty, journal fit, every figure and table) using independent review agents.
6. Stay with MIMIC-IV only; revise the paper to a high standard (new analyses on existing data: data-scarce sites, care-era partition, stealthier attacks, calibration fixes, unified statistics; reframing; trimming; reference corrections).
7. Select a target journal (the authors chose Blockchain in Healthcare Today) and format the manuscript to its requirements.

#### What the assistant was told not to do

Fabricate results or references; share or commit patient-level data or model predictions (PhysioNet data use agreement); overstate novelty; hide negative results.
