## Supplementary material for "A Blockchain-Enabled Federated Ensemble Framework for Privacy-Preserving Mortality Prediction Across Heterogeneous Healthcare Systems": Tripod

### Supplementary File S3. TRIPOD+AI checklist and PROBAST risk-of-bias notes

Locations are given as section numbers of the manuscript (final build of 19 November 2026), which are the same in the Word and PDF versions. If the journal asks for the official TRIPOD+AI form (tripod-statement.org), transfer these locations onto it.

#### A. TRIPOD+AI (Collins et al., BMJ 2024)

| Item | Topic | Location | Status |
| --- | --- | --- | --- |
| 1 | Title | Title | Reported: prediction, setting, simulated consortia |
| 2 | Abstract | Abstract | Reported (structured, BHTY format) |
| 3a–c | Background: context, rationale, existing models | §1, §2.1–2.2 | Reported |
| 4 | Objectives | §1, Abstract | Reported |
| 5a–b | Data source, dates | §3.1 | Reported (MIMIC-IV v3.1, 2008–2022) |
| 6a–c | Participants: setting, eligibility, treatments | §3.1; Supplementary Fig. S1 | Reported |
| 7 | Data preparation | §3.1–3.2; Supplementary Methods S-M4 | Reported |
| 8a–c | Outcome: definition, timing, blinding | §3.1 | Reported (in-hospital death; 24 h landmark); blinding not applicable |
| 9a–c | Predictors: selection, definition, timing | §3.1; Supplementary Table S1; S-M1 (extended sets) | Reported |
| 10 | Sample size | §3.1, §4.1 | Partly: whole available cohort (9,984 events); no formal calculation |
| 11 | Missing data | §3.1; S-M4 | Reported (informative missingness; native handling in trees; imputation with indicators for MLPs) |
| 12a–g | Analytical methods | §3.2–3.8; use of AI tools §3.9 | Reported |
| 13 | Class imbalance | §3.8 | Reported (no resampling; AUPRC, calibration and threshold metrics reported) |
| 14 | Fairness | §3.8, §4.4 (A6), §4.6; Figs 5–6; Supplementary Tables S7–S8 | Reported |
| 15 | Model output, thresholds | §3.5 | Reported (federated recalibration; F1-optimal federated threshold) |
| 16 | Development vs evaluation data | §3.2, §5.5 | Reported |
| 17 | Ethical approval | Ethics | Reported |
| 18a–f | Funding, conflicts, protocol, registration, data, code | Funding, Disclosures, Data availability, Code availability; Reporting | Reported; not registered (stated); code on GitHub and Zenodo (link in the cover letter and the medRxiv preprint; removed from the blinded manuscript) |
| 19 | Patient and public involvement | Ethics | Reported (none) |
| 20a–c | Participant flow, characteristics, events | §3.1, §4.1; Table 2; Supplementary Fig. S1, Table S2 | Reported |
| 21 | Numbers of participants and events in development | §4.1; Table 2 | Reported |
| 22 | Model specification | §3.3–3.5; S-M4; code release | Reported |
| 23a–b | Performance, incl. by site and subgroup | §4.2–4.8; Tables 3–8; Figs 2–6; Supplementary Tables S3–S11 | Reported |
| 24 | Model updating | — | Not applicable (local intercept update evaluated as a variant, §4.5) |
| 25 | Interpretation | §5.1–5.4 | Reported |
| 26 | Limitations | §5.5 | Reported |
| 27a–c | Usability, data quality, future research | §5.2, §5.4, §6 | Partly: no implementation study |

#### B. PROBAST notes (Wolff et al., Ann Intern Med 2019; applied informally to a development study with internal evaluation)

| Domain | Judgement | Reasons |
| --- | --- | --- |
| Participants | Low concern for bias; high concern for applicability | One academic centre (MIMIC-IV); appropriate inclusion (adults, stay ≥24 h, alive at 24 h). Hospitals were simulated from one institution, so transfer to real multi-institution consortia is untested. |
| Predictors | Low | Predictors restricted to data available at 24 h; comorbidities from previous admissions only, avoiding discharge-coding leakage; definitions fixed before modelling. |
| Outcome | Low | In-hospital death recorded in the database, objective and not influenced by predictors. |
| Analysis | Some concerns | Large number of events (9,984). Patient-level splits; clustered bootstrap for inference; calibration reported pooled and by site. Concerns: validation splits were reused for early stopping, attestation, recalibration and threshold selection; several analyses were added after first results; no external validation. |
| Overall | Some concerns; applicability limited to simulated consortia from one centre | As stated in §5.5. |
